# Widespread recessive effects on common diseases in a cohort of 44,000 British Pakistanis and Bangladeshis with high autozygosity

**DOI:** 10.1101/2024.04.03.24305256

**Authors:** Teng Hiang Heng, Klaudia Walter, Qin Qin Huang, Juha Karjalainen, Mark J Daly, Henrike Heyne, FinnGen, Daniel Malawsky, Georgios Kalantzis, Genes & Health Research Team, David A. van Heel, Hilary C Martin

## Abstract

Genetic association studies have focused on testing additive models in cohorts with European ancestry. Little is known about recessive effects on common diseases, specifically for non-European ancestry. Genes & Health is a cohort of British Pakistani and Bangladeshi individuals with elevated rates of consanguinity and endogamy, making it suitable to study recessive effects. We imputed variants into 44,190 genotyped individuals, using two imputation panels: a set of 4,982 whole-exome-sequences from within the cohort, and the TOPMed-r2 panel. We performed association testing with 898 diseases from electronic health records. We identified 185 independent loci that reached standard genome-wide significance (p<5×10^−8^) under the recessive model and had p-values more significant than under the additive model. 140 loci demonstrated nominally-significant (p<0.05) dominance deviation p-values, confirming a recessive association pattern. Sixteen loci in three clusters were significant at a Bonferroni threshold accounting for multiple phenotypes tested (p<5.5×10^−12^). In FinnGen, we replicated 44% of the expected number of Bonferroni-significant loci we were powered to replicate, at least one from each cluster, including an intronic variant in *PNPLA3* (rs66812091) and non-alcoholic fatty liver disease, a previously reported additive association. We present novel evidence suggesting that the association is recessive instead (OR=1.3, recessive p=2×10^−12^, additive p=2×10^−11^, dominance deviation p=3×10^−2^, FinnGen recessive OR=1.3 and p=6×10^−12^). We identified a novel protective recessive association between a missense variant in *SGLT4* (rs61746559), a sodium-glucose transporter with a possible role in the renin-angiotensin-aldosterone system, and hypertension (OR=0.2, p=3×10^−8^, dominance deviation p=7×10^−6^). These results motivate interrogating recessive effects on common diseases more widely.

**Graphical Abstract:** 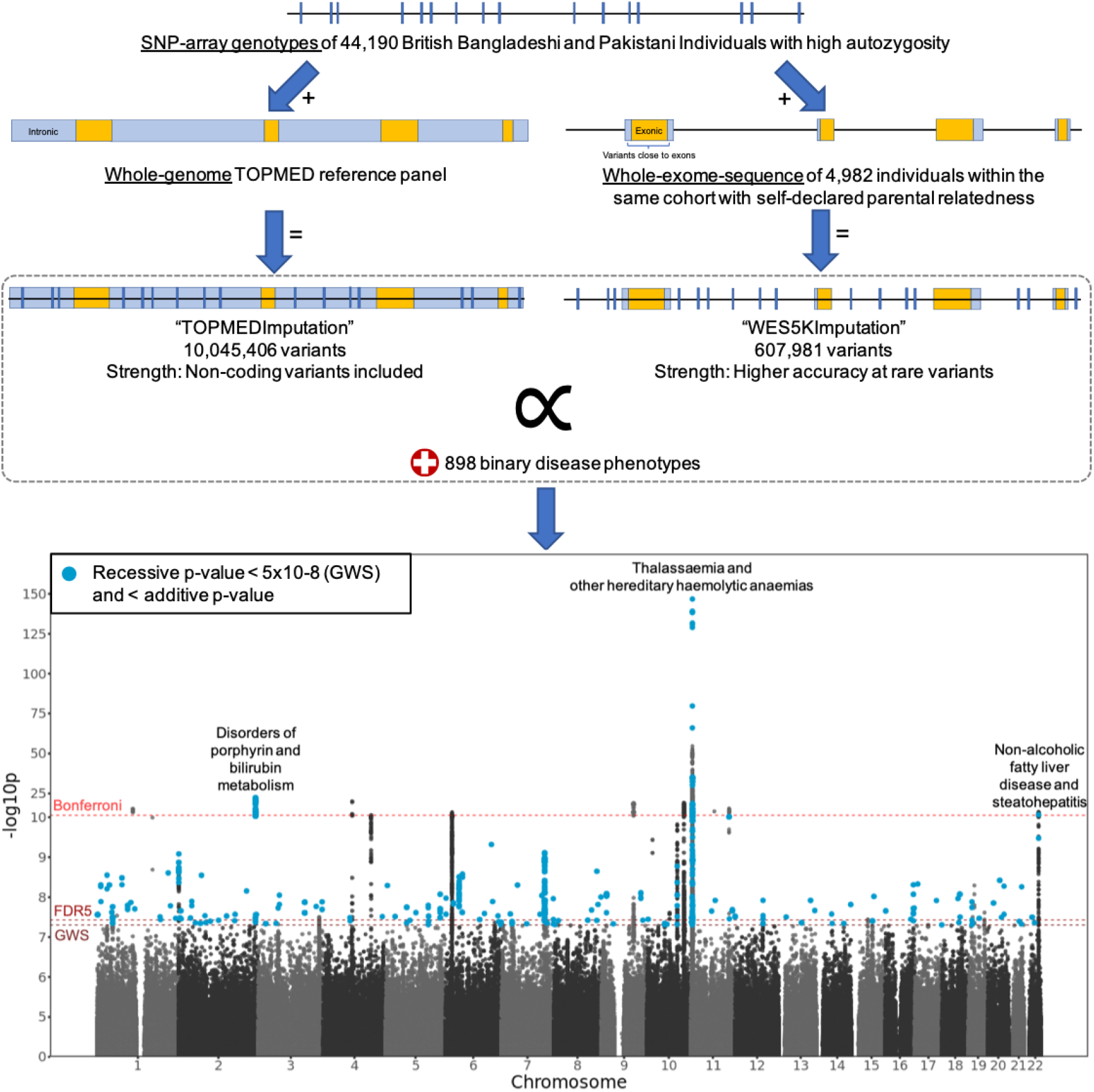

## Introduction

Recessive effects in humans have been primarily studied in the context of rare, monogenic disorders, and little is known about recessiveness in common diseases and complex traits ^1^. Identifying variants with recessive associations to diseases could improve polygenic risk scoring, provide better insight into gene and variant function, improve understanding of disease pathophysiology, and allow for the identification of novel drug targets ^2–4^.

The effects of genetic variation on common complex phenotypes are typically discovered through genome wide association studies (GWASes) ^5–7^, where an additive model is predominantly tested. However, applying a recessive model has allowed for the discovery of associations that would have been otherwise missed under conventional additive testing. For example, Heyne et al. (2023) ^8^ performed recessive tests on 44,370 variants and 2,444 diseases in the FinnGen project from Finland, and identified thirty-one loci at genome-wide significance (GWS, p<5×10^−8^) where the associations were more significant in the recessive model than in the additive model. Of the twenty findings further validated, notably thirteen loci would have been missed with the additive model alone. Similarly Guindo-Martinez et al. (2021) ^9^ performed non-additive association testing in 62,281 subjects across twenty-two age-related diseases, and amongst twenty-six novel loci, four were identified only with the recessive model. Palmer et al. (2023) ^1^ systematically quantified the contribution of dominance deviations (deviation from the additive pattern of inheritance) to heritability across 1,060 common traits in UK Biobank (n = 361,194). They identified non-additive effects of 183 phenotype-locus pairs across the phenotypic spectrum, but concluded that overall, non-additive effects contribute very little to heritability. Collectively, these suggest that many recessive associations on common traits still remain to be found through the application of non-additive testing, including with rare variants, where the additive model would be more likely to miss recessive effects.

With the construction of large-scale biobanks, it is now possible to study the recessive contribution to common diseases ^8,10^. However, in outbred populations, very large sample sizes are required for adequate power to test recessive effects than additive effects, particularly for rare variants. Power to detect recessive effects is expected to be increased in bottlenecked populations like Finland, where recessive variants may rise in frequency due to founder effects ^8,11^, or in populations enriched for consanguinity and therefore, increased homozygosity. We showed in simulations that power to find recessive effects is boosted both by explicitly testing a recessive model and by increased homozygosity (Appendix-1).

Genes & Health (G&H) is a community-based cohort of currently ∼60,000 individuals of British Bangladeshi and Pakistani ancestry with genetic data and linked electronic health records (EHRs) ^12^. The cohort has a high rate of consanguinity (32% are offspring of second cousins or closer). Furthermore, British Pakistanis, who comprise 40% of the cohort, have been previously found to have high levels of endogamy, due to the biraderi system, with multiple bottlenecked subpopulations displaying elevated levels of identity-by-descent (IBD) sharing, more than ten to twenty times than found in the Finnish ^13,14^. We therefore hypothesised that the increased IBD-sharing within these subgroups in G&H might allow reasonable quality imputation of variants even from a relatively small sample size of individuals from the same cohort. From this imputation, we would be able to take advantage of the increased homozygosity to test for recessive effects.

We leveraged genotype chip data on 44,000 G&H individuals, of whom around 5,000 also had whole-exome sequencing (WES) data. Firstly, we imputed variants from the exome-sequenced individuals into the larger genotyped G&H cohort. This was inspired by Barton et al. (2021) ^15^, who boosted power for association testing of rare coding variants by building a within-cohort reference panel from 49,960 WES samples in UK Biobank and imputing variants into the larger genotyped cohort (n ∼ 500,000). We also used an additional imputation reference panel, the Trans-Omics for Precision Medicine (TOPMED-r2) panel (97,256 individuals including 644 with South Asian ancestry) ^16^ to perform whole-genome imputation to study recessive effects in the noncoding regions. Using these two imputed datasets, we performed association testing with binary phenotypes curated from the electronic health records, focusing on detecting recessive effects.

## Methods

### Preparation of genetic data

#### Preparation of the genotyped data

Detailed quality control (QC) of the genetic data is described in Appendix-3. Genome-wide genotyping was performed with the Illumina Global Screening Array (GSAv3EAMD, build 38), and these data were used as the imputation backbone. Initial QC has been described in Huang et al. (2022) ^17^. From the genotyped 44,396 individuals, we inferred 44,190 individuals to be either of Bangladeshi or Pakistani genetic ancestry (Appendix-4), and downstream analyses were restricted to these individuals. Another round of variant filters was applied to include only autosomal, bi-alleleic SNPs with ≥99% call rate. The Pakistani subgroup has high autozygosity and strong population structure, while the Bangladeshi subgroup has minimal structure and much less autozygosity ^18^, so to avoid excluding too many high-quality variants due to failure on a standard test for Hardy-Weinberg Equilibrium (HWE), the HWE test was performed (using PLINK1.9) ^19^ only in the Bangladeshi subgroup and the variants that failed a p-value threshold of 10^-6^ in Bangladeshis were then excluded from the entire dataset. Variants with MAF>0.1% were included from the imputation backbone, which resulted in 469,678 variants that were then phased with EAGLE2 (Kpbwt=20,000) ^20^.

#### Preparation of the within-cohort imputation panel

Exome sequencing was performed using Agilent V5 capture kits on a subset of 5,236 individuals who self-declared as having consanguineous parents.

Mapping, calling and initial QC included excluding samples with sex discrepancies and <10X on-target coverage, and applying the following variant filters using bcftools ^21^: “QD < 2.0 || FS > 30 || MQ < 40.0 || MQRankSum < -12.5 || ReadPosRankSum < -8.0” for SNPs and “QD < 2.0 || FS > 30 || ReadPosRankSum < -20.0” for indels ^22^. We subsetted to the 5,073 individuals who were genetically inferred to be of Bangladeshi or Pakistani ancestry from their array data. Then we set to missing genotypes that had genotype quality (GQ) <20, a p-value from a binomial test for allele depth at heterozygous sites (binomAD) <10^-2^, or depth ≤7. Variants were excluded if they had a post-genotype QC call rate <70%. 91 samples with high missingness or high discordance with their array data were excluded, leaving 4,982 samples (Appendix-5). The WES data were then merged with the SNP-array data and phased with EAGLE2 (Kpbwt=20,000) to form a reference panel for imputation.

#### Imputation

We imputed the G&H data against two different imputation panels. Firstly, variants were imputed from the within-cohort whole-exome reference panel (described above) into the individuals without WES data with Minimac4 ^23^. To assess the imputation accuracy and determine an imputed R^2^ cutoff, ten “leave-10%-out” trials were performed (Appendix-5). We retained variants with imputed R^2^ ≥0.5 and at least three individuals with a homozygous genotype (N_Hom_ ≥3). This is referred to as the “WES5Kimputation” dataset. The SNP-array data were also submitted to the TOPMed-r2 Minimac4 1.5.7 Imputation Server ^16,23,24^ for whole-genome imputation against the TOPMED-r2 panel. The same post-imputation filters were applied, and this is referred to as the “TOPMEDimputation”.

#### Variant annotation

Variants were annotated with Ensembl’s Variant Effect Predictor (VEP) v107. For each variant, the worst consequence for any transcript was extracted for subsequent analyses.

### Phenotype curation

Two lists of phenotypes were curated from participants’ electronic health records, and phenotype information was encoded as a binary with ‘1’ coding for a case and ‘0’ coding for a control. A list of 237 custom phenotypes were compiled manually, and a second set of 1,281 phenotypes were defined based on International Classification of Disease (ICD10) codes. Further detail is described in Malawsky et al. (2023) (Methods section on “Phenotypic data harmonisation and preparation for G&H”) ^18^. We retained phenotypes with ≥30 cases and also classified them into those that affected both sexes or were sex-specific (i.e. occurred only in females or males). For sex-specific phenotypes, the cohort was filtered to the relevant sex for testing. This resulted in 898 phenotypes.

### Association testing

Association testing was performed using REGENIE ^25^ through its two-step pipeline. In step 1, the model was fitted using variants from the SNP-array, using the leave-one-out cross-validation (LOOCV) scheme and a genotype block size of 1,000. Step 2 was performed under the additive model and the recessive model, at a genotype block size of 1,000 and p-value threshold of 0.05, below which the approximate Firth correction was applied. The covariates included age, sex, age^2^, age x sex, age^2^ x sex and the first ten principal components (PCs) from the principal component analysis on unrelated G&H individuals (Appendix-4).

We defined significant recessive associations as tests with p-values <5×10^−8^ (GWS), and with a p-value lower under the recessive model than under the additive model. We excluded the human leukocyte antigen (HLA) region due to complex LD (chr6:25mb-35mb), and then defined independent loci with the following steps: (1) For each phenotype with significant tests, we identified the test with the most significant recessive p-value as the lead variant. (2) We calculated the LD R^2^ using PLINK1.9 between the lead variant and variants within a +/- 1.5Mb window of it. Variants with an LD R^2^ ≥0.25 with the lead variant were defined as being part of the same locus, following previous work from FinnGen^8,11^. (3) We then identified the next most significant variant amongst the remaining variants that are not part of the locus defined in the previous steps, and repeated the loop.

We calculated dominance deviation p-values to assess the evidence that our significant associations detected under the recessive model were really recessive. Specifically, for the lead variants of the significant recessive associations, we ran logistic regression in R, controlling for the same covariates, performing genotypic tests with 2 degrees of freedom as well as additive and recessive tests for comparison. Description of how this was performed and detailed comparisons of the results with those from REGENIE are described in Appendix-9.

### Testing for replication in FinnGen and GERA

FinnGen is a public-private collaboration to profile the genomic and digital healthcare data of ∼500,000 Finnish individuals, with the goal to uncover novel biological and therapeutic insights into human diseases. As a recessive testing pipeline has been built in the cohort before ^8^, we used it as an independent cohort to assess replication. More details of the cohort are described in Appendix-12. Phenotypes were matched manually to FinnGen binary phenotypes curated from Finnish health registers (Table S7). The variants from FinnGen release 10 were then tested with the phenotypes using REGENIE under the recessive model, controlling for the covariates sex, age, 10 PCs and genotyping batch, as described for release 10 on (https://www.finngen.fi/). ^11^ For each significant locus in G&H, we first identified proxy variants as variants that are within the window described above (+/- 1.5Mb from the lead variant with LD R^2^ ≥0.25. For a more stringent assessment of replicability, we applied the locus definitions from Huang et al.^17^ and repeated the analysis (see Appendix-10). We defined a locus as replicating (or ‘transferable’) in FinnGen if any of the lead or proxy variants had the same direction of effect as observed in G&H at a nominally-significant p-value <0.05, when tested in a recessive model with a similar trait. To compare the number of loci that replicated to what we might have expected to replicate given the power in FinnGen, we calculated a power-adjusted transferability ratio ^17^. Specifically, for each locus, we used the effect size of the lead variant in G&H, the allele frequency of the lead variant in FinnGen, and the case rate and sample size in FinnGen to estimate the power for the test with the genpwr R package ^26^. The expected number of transferable loci was calculated as the sum of the power estimates across all the loci. The power-adjusted transferability (PAT) ratio was then calculated by dividing the observed number of transferred loci by the expected number.

Publicly-available summary statistics from recessive testing in the Genetic Epidemiology Research on Ageing (GERA) cohort by Guindo-Martinez et al. (2021) ^9^ were accessed on 14 December 2023. Phenotypes were matched manually (Table S8) and the PAT ratio was calculated as described above, using allele frequencies and case rates in GERA.

## Results

### Recessive association testing identifies 185 loci

Using exome sequence data from 4,982 individuals from Genes & Health, we generated a reference panel which allowed us to impute 605,263 variants, including rare exonic variants, into the larger cohort of 44,186 individuals from that cohort. Simultaneously, we carried out a whole-genome imputation of 10,045,406 variants using the TOPMED panel. We then tested these variants for recessive associations with 898 phenotypes. We identified 185 unique loci where the lead variant had a genome-wide significant recessive p-value (<5×10^−8^) that is smaller than the additive p-value (Figure-1, Table S5). At a stringent Bonferroni cutoff (0.05/9,197,933,046 tests = 5.5×10^−12^, Appendix-6), 16 loci remained. 144 loci passed a more lenient Benjamini–Hochberg cutoff for a false discovery rate of 5% (FDR 5%) (p< 3.7×10^−8^, so close to the genome-wide significance threshold). Notably, the sixteen Bonferroni-significant findings can be found in three clusters (Figure 1; Table 1), corresponding to (a) Non-alcoholic fatty liver disease and steatohepatitis (NAFLD) (one locus; lead SNP chr22:43939790), (b) disorders of porphyrin and bilirubin metabolism (one locus, found with both imputation panels; lead SNP chr2:233763993), and (c) thalassaemia and other hereditary haemolytic anaemias (13 loci, one of which was found with both imputation panels; lead SNPs at chr11:4908482-5544800).

**Figure 1.**
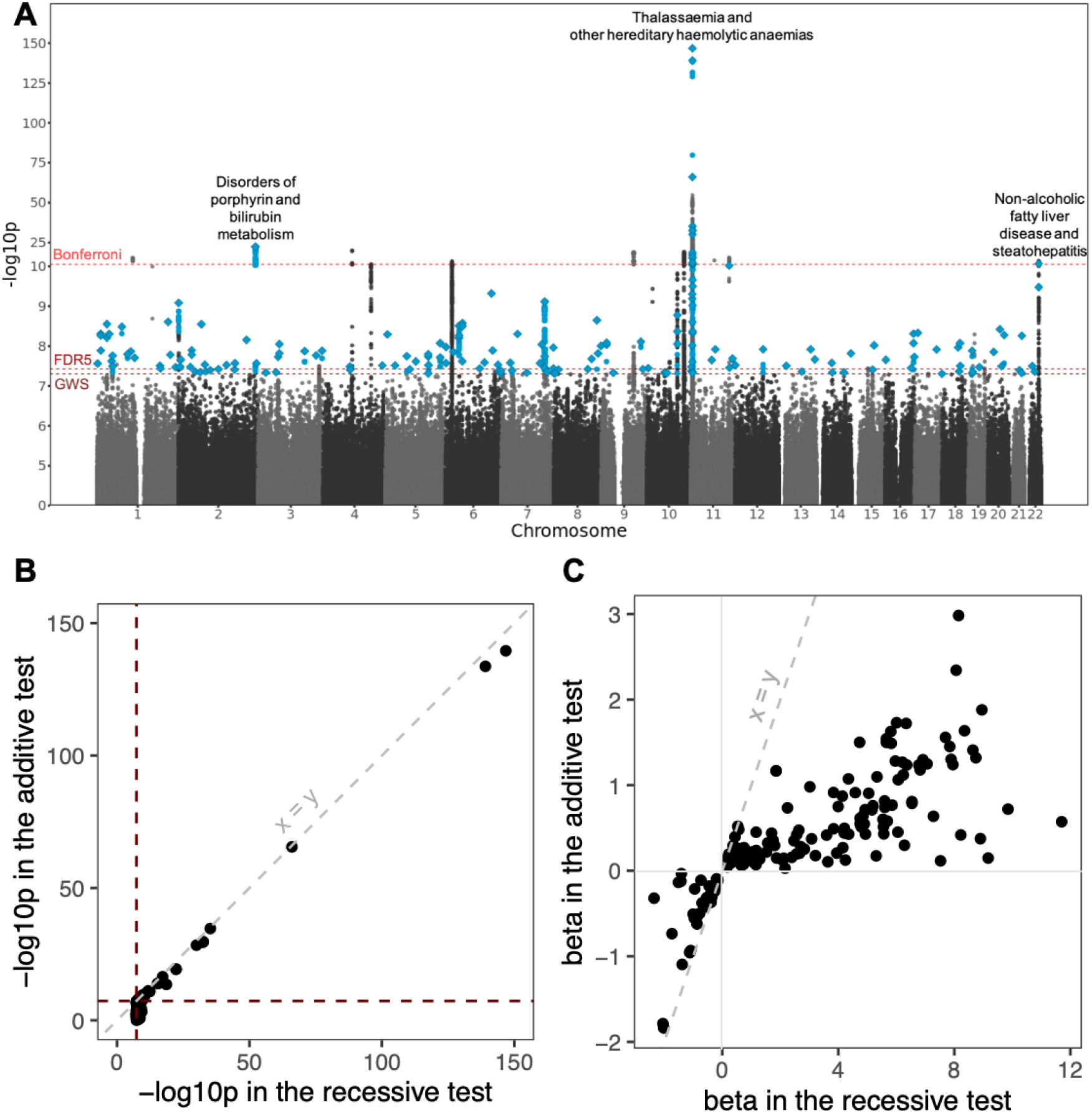
The 185 loci identified at genome-wide significance in recessive association tests. A: Manhattan Plot of the results from recessive tests performed. Dashed horizontal lines represent the various p-value cutoffs: genome-wide significance (p<5×10^−8^, “GWS”), for a false discovery rate of 5% (p<3.7×10^−8^, “FDR5”), and the Bonferroni-corrected cutoff (p<5.5×10^−12^, “Bonferroni”). Blue points represent recessive tests passing the genome-wide significance threshold where the recessive p-value is more significant than the additive p-value, and diamonds represent lead variants defined as described in Methods. The clusters containing Bonferroni-significant associations are labelled with the phenotypes with which they are associated. B: For the lead variants, the -log10p from additive testing against -log10p from recessive testing. The horizontal line indicates the genome-wide significance threshold, and the diagonal line is y=x. C: The effect size (beta) of the genotype encoding from additive testing against the beta from recessive testing for the lead variants.

**Table 1.**
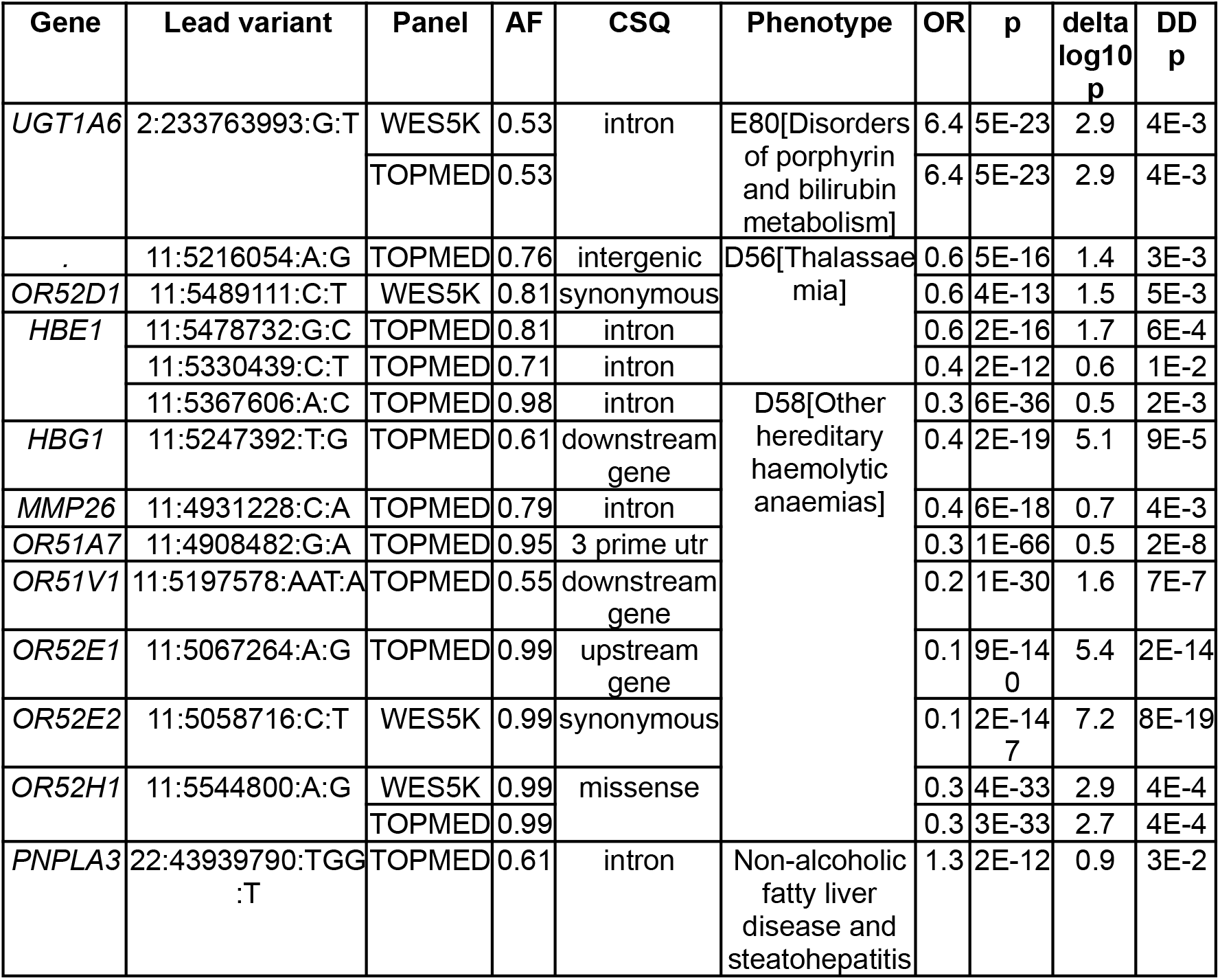
Table of recessive findings that passed Bonferroni significance. “CSQ”: consequence annotation of the variant. “OR”: Odds ratio, converted from beta in the test output. “delta log10p”: The difference between the -log10p in the recessive test and the -log10p in the additive test. “DD p”: Dominance deviation p-value. The allele frequence (AF) and odds ratio (OR) presented are for the alternate allele (second allele in the lead variant ID).

Of the lead variants at the 185 recessive loci, 152 (82%) were not genome-wide significant in the additive test (Figure-1B), suggesting that they would have been missed in conventional additive GWAS-es. The associations that were not genome-wide significant in the additive test tended to be with rarer variants (Figure-S14A). The effect sizes in the recessive tests for these lead variants also tend to have a larger magnitude than in the additive tests (Figure-1C).

Seventy-six percent (140) of these lead variants had nominally significant dominance deviation p-values.

These 185 recessive loci include loci that were significant in either or both of the imputation panels. Twenty-nine of the lead variants were successfully tested in both imputation sets. At these, effect sizes (Figure-2B) and allele frequencies (Figure-2C) correlated well between the two imputation panels, though interestingly, a small subset of findings significant with the WES5Kimputation had much less significant p-values when their TOPMEDimputation genotypes were tested (Figure-2A). These findings corresponded to variants with lower imputed R^2^ values in the TOPMEDimputation compared to the WES5Kimputation (Figure-2D), suggesting that higher confidence in the imputation when using the in-house panel improved the sensitivity of the testing through more accurate prediction of the genotypes.

**Figure 2.**
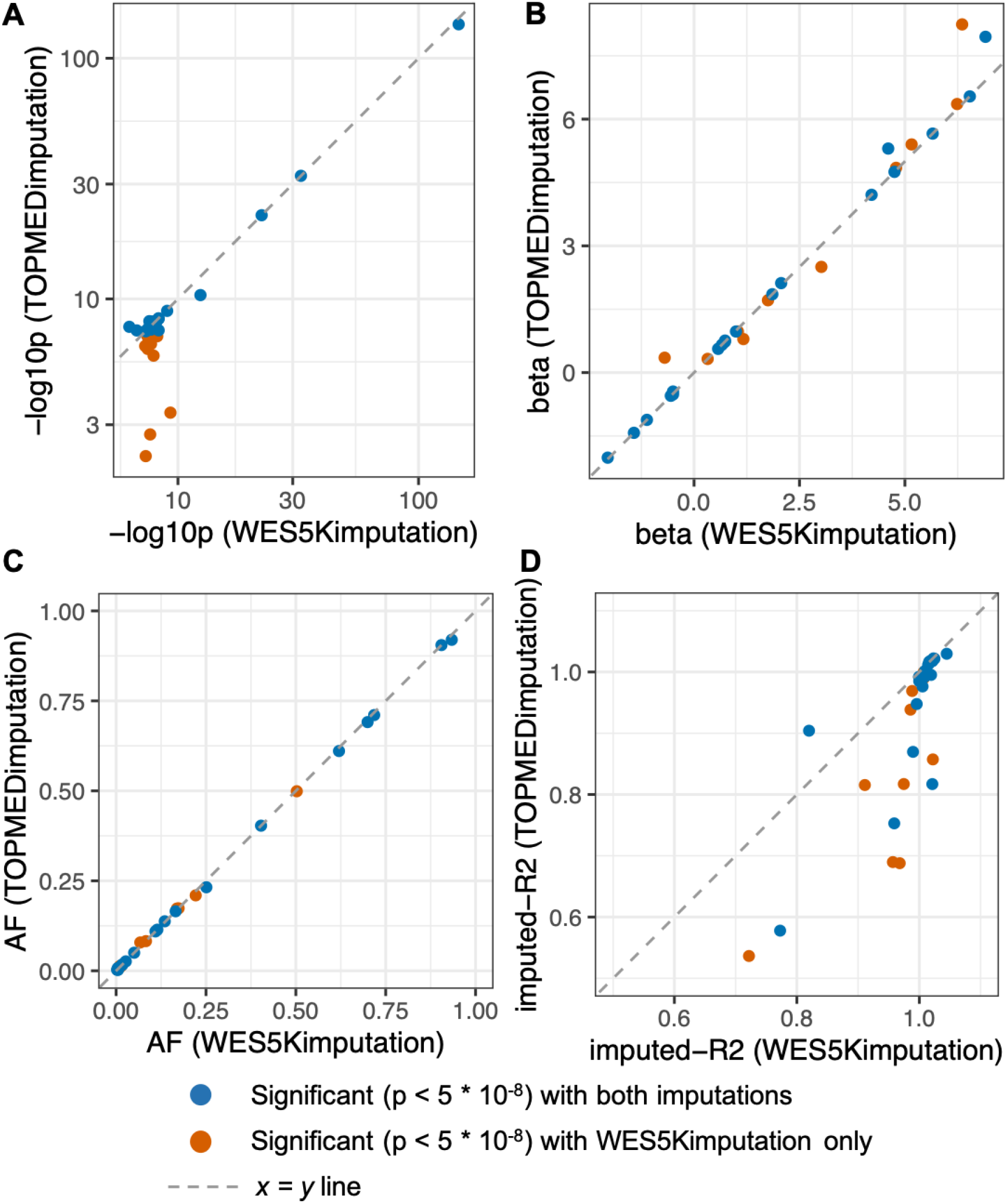
Comparison of results for variants tested using both imputation panels and significant in one or both. Scatter plots show the (A) p-values (-log10p), (B) betas, (C) allele frequencies (AFs), (D) imputed-R^2^ values of lead variants.

### Replication of significant findings in other cohorts

We sought to test how many of the 185 loci could be replicated in two other cohorts suitable for recessive association testing, FinnGen and GERA. 133 loci could be found in FinnGen, meaning the lead variant of the locus could be tested with a comparable phenotype in FinnGen. Of these 133 lead variants, 82 had the same direction of effect in the recessive tests of both cohorts. This is significantly more than the number expected by chance (one-sided exact binomial test, p-value 0.0045).

Next, we calculated the power-adjusted transferability (PAT) ratio (Huang et al. 2022), a quantitative measure of replicability, measuring how many loci can be replicated in FinnGen relative to the power in this independent cohort, taking into account allele frequencies, case rates and sample sizes. We expected to replicate 129 of the 133 examined loci at p < 0.05; of these, 30 loci replicated in FinnGen, resulting in a power-adjusted transferability ratio of 28%. This suggests that a p<5×10^−8^ cutoff for significance in Genes & Health is too lenient given the number of phenotypes tested. Restricting to the Bonferroni-significant findings, the power-adjusted transferability ratio increased to 44%. The loci associated with NAFLD and disorders of porphyrin and bilirubin both replicated. For the cluster of loci associated with thalassemia and other hereditary haemolytic anaemia, four out of thirteen loci replicated, while eight of the remaining nine loci had the same direction of effect in both cohorts (although we note that we were not considering identical phenotypes in FinnGen since they were not available - see Table S7).

We also assessed replication in the GERA cohort. The twenty-two phenotypes tested by GERA restricted the number of phenotypes we could match, so only eleven GWS loci could be evaluated. After accounting for AF, case rate and sample size differences, we expected to replicate around eight loci. Three loci replicated, resulting in a power-adjusted transferability ratio of 36%.

### The Bonferroni-significant recessive associations

The Bonferroni-significant lead variants are listed in Table 1. We summarise these significant associations below.

We found a recessive association between rs66812091 (chr22:43939790:TGG:T), an intronic variant in the gene *PNPLA3* (Patatin-like Phospholipase Domain-containing Protein 3), and NAFLD, with an odds ratio of 1.3 at a p-value of 2×10^−12^. This p-value is an order of magnitude more significant in the recessive test than the additive test (2×10^−11^). The relationship also had a significant dominance deviation p-value of 3×10^−2^ and was replicated in FinnGen with a recessive test OR of 1.3 and p-value of 6×10^−12^. *PNPLA3* is involved in lipid and fatty acid metabolism and has high expression in adipose and liver tissues ^27,28^. This gene’s role in fatty acid metabolism and liver function has been further supported by functional knockout studies in mice ^28,29^. This variant has also been shown in additive GWASes to be associated with NAFLD ^30^ and deranged liver enzymes (a marker of hepatitis) ^31^. However, to our knowledge, this is the first study demonstrating that in fact one or more of the variants near this gene are likely to have recessive rather than additive effects on NAFLD.

Next, an intronic SNP in the gene *UGT1A6*, rs6742078 (chr2:233763993:G:T), identified with both imputation panels, was found to be associated with disorders of porphyrin and bilirubin metabolism. *UGT1A6* encodes a uridine 5’-diphospho-glucuronosyltransferase (UDP-glucuronosyltransferase, UGT). UGTs are enzymes responsible for the glucuronidation of lipophilic molecules, including bilirubin, into the conjugated, hydrophilic form that can be excreted in the urine ^32^. *UGT1A6* has been shown to be associated with bilirubin levels in the GWASs on liver traits and bilirubin levels^33,34^. Mutations in UGTs, specifically *UGT1A1*, cause the well-known autosomal recessive conditions Gilbert’s syndrome ^35^ and Crigler-Najjar syndrome ^35,36^, in which patients present with excess unconjugated bilirubin. The findings presented here demonstrate novel recessive associations between another UGT not previously reported and dysfunctional bilirubin clearance.

Thirdly, we found multiple Bonferroni-significant recessive hits in the genomic region chr11:4908482-5544800 associated with thalassaemia and other hereditary haemolytic anaemias. These variants are also associated with multiple anaemia traits in additive GWASs ^37,38^. They span two haemoglobin genes, haemoglobin epsilon locus (*HBE1*) and haemoglobin gamma a (*HGB1*), as well as the gene *MMP26* and a cluster of olfactory receptor genes. The two haemoglobin genes *HBE1* and *HGB1* encode for embryonic and foetal haemoglobin (HbF) subunits respectively. The gene *HBG1* is well established to be associated with beta-thalassemia and haemoglobin E disease ^39^.

Beta-thalassaemia is an inherited anaemia caused by dysfunctional production of the beta globin subunit of adult haemoglobin (HbA) ^40^, therefore, persistence of elevated HbF past infancy as a compensatory mechanism is occasionally observed ^41^. Genes encoding embryonic haemoglobin and HbF, including *HBE1* and *HGB1*, have been targeted as part of possible therapies in thalassaemias, where reactivating these alternative forms of haemoglobin expression may help to supplement low HbA production ^42–44^. The variants that we find to be associated with thalassaemia and anaemias near these genes may be involved in the regulation of haemoglobin expression and inactivation.

### Recessive findings implicating coding variants provided insight into gene functions

We also found several recessive associations for coding variants that passed our more lenient p-value threshold of 5×10^−8^. Since these coding variants seem likely to be the causal variants at those loci, we discuss several examples here.

Firstly, we see a recessive association between a missense variant (chr1:11796321:G:A, rs1801133) in methylenetetrahydrofolate reductase (*MTHFR*) and folate deficiency (OR = 2.1, p-value = 5×10^−9^, dominance deviation p-value = 10^-3^). This finding was also replicated in FinnGen (OR = 2.1 p-value = 0.02). *MTHFR* is an enzyme involved in folate and homocysteine metabolism. After folate is converted to 5,10-methylenetetrahydrofolate (5,10-MTHF), *MTHFR* reduces 5,10-MTHF to 5-methyltetrahydrofolate (5-MTHF), which is then required as a cosubstrate for the conversion of homocysteine to methionine ^45^. The missense variant we report (C677T) has been shown to cause instability in *MTHFR*, resulting in the accumulation of homocysteine ^46^. Indeed, there are reports that this C677T mutation has a recessive effect on homocysteine levels, but with a mild heterozygous effect: individuals heterozygous for the C677T mutation have mildly elevated homocysteine levels, while the homozygous individuals have significantly higher levels ^47–49^. Functional studies have shown that the enzyme’s reduced efficiency from instability caused by the C677T transition can be compensated with additional folate ^48,50–52^. This would result in low folate levels, providing a possible biological explanation for our association. Multiple additive GWASes have reported associations between this variant and folate deficiency anaemia ^30^ or being on folate supplements ^53^. However, our study suggests that the underlying pattern of inheritance may be recessive instead. Folate is essential for DNA, RNA and protein methylation, and foetal neural tube defect is an established consequence of low folate levels in pregnancy ^54^. Germane to this, there are several reports (although conflicting) of recessive associations between variants (including C677T) in *MTHFR* and neural tube defects ^55–58^.

As a second example, we have found a protective recessive association between a missense variant in *SLC5A9* (chr1:48228922:G:A, rs61746559) hypertension (OR = 0.2, p-value = 3×10^−8^, dominance deviation p-value = 7×10^−6^). *SLC5A9*, solute carrier family 5 member 9, is also known as sodium-glucose transporter 4 (*SGLT4*), and is a member of the solute carrier (SLC) superfamily. Specifically, it is a sodium-dependent glucose transporter of mannose, 1,5-anhydro-D-glucitol, and fructose ^59^. Another member of this family is the well-studied sodium-glucose transporter 2 (*SGLT2*), a glucose transporter largely expressed in the kidney, and is a target of gliofozins, drugs used for lowering serum glucose levels in patients with type 2 diabetes ^60^. It has been shown that a concomitant benefit of inhibiting *SGLT2* in type 2 diabetes patients is lowering blood pressure ^61–63^, possibly from haemodynamic changes in kidney glomeruli ^64^ and regulating the renin–angiotensin–aldosterone system ^65^. *SGLT4* may function in a similar manner, as, like *SGTL2*, it is expressed in the kidneys ^59^. Missense mutations may inhibit its function, explaining the relationship we detect with hypertension. While there is limited information about this gene’s function currently ^66^ to support this hypothesis, there is an additive association between this variant and renin levels ^67^, suggesting it may indeed be involved in the renin–angiotensin–aldosterone system. Multiple widely-used classes of antihypertensives work on the renin-angiotensin-aldosterone system, which plays a key role in regulating blood pressure^68^.

## Discussion

After performing recessive association testing on variants imputed from both a within-cohort exonic reference panel and the whole-genome TOPMED reference panel, 185 unique loci were identified at genome-wide-significance, where the recessive association was more significant than the additive association. After Bonferroni multiple testing correction, sixteen loci in three clusters remained. After adjusting for changes in case rates, allele frequencies and sample size in the independent cohort FinnGen, we replicated 44% of the expected number of Bonferroni-significant loci we would be powered to replicate, including at least one locus from each cluster. We also identified recessive associations at loci previously thought to be additive. Examples include the association between rs66812091 and NAFLD, and between rs1801133 and folate deficiency. Notably, we report a novel recessive association between a missense variant in *SGLT2* (rs61746559) and reduced risk of hypertension.

In modelling binary traits, one usually assumes a liability threshold model, in which an individual develops the disease once they pass a certain threshold on a continuous, quantitative trait (the ‘liability’) that follows a normal distribution in the population ^69^. Under this model, it is possible that a variant may have an additive effect on the underlying liability but a recessive effect on disease status. For example, it may be that rs66812091 in *PNPLA3* has an additive effect on liver enzyme levels, leading to the accumulation of fatty acids that results in hepatic inflammation (i.e. the underlying quantitative trait), but a recessive effect on NAFLD (p = 2.4×10^−12^). Supporting this, Barton et al. (2022)^70^ showed in UKBB that variants with known recessive associations to disease can have milder heterozygous effects in related quantitative traits.

Therefore, characterising the underlying inheritance pattern in greater detail could improve our understanding of disease pathophysiology and highlight homozygous individuals as higher-risk groups to target during screening.

With G&H, we were able to perform imputation with a within-cohort reference panel, and demonstrated the advantage of using this to improve sensitivity, as previously reported by Barton et al. (2021) ^15^. We saw a subset of recessive findings with p-values that were only significant with the WES5Kimputation and not the TOPMEDimputation. We hypothesise from our evaluation of imputation accuracy (Appendix-5) that this might be due to more accurate imputation of these variants with the within-population reference panel. However, the TOPMEDimputation provided a significantly larger set of variants to work with, which could likely explain why most of our associations are from that panel.

Our recessive testing highlights the value of detecting novel recessive associations that would have been missed under the additive model. For example, the rs61746559 missense variant affecting SGLT2 was previously reported to be additively associated with renin levels, and with the novel finding that the homozygous genotype may be protective for hypertension, it may provide a basis to consider the gene as a drug target. The majority of the GWS findings (152 / 185) were not GWS under the additive model, illustrating the importance of applying the recessive model of testing for finding recessive effects. For example, we showed that an intronic variant (chr13:109179501:G:C, rs2038707) in myosin 16 (*MYO16*), a gene involved in the musculoskeletal system ^71,72^, had a recessive association with the ICD10 code M85 “Other disorders of bone density and structure” (OR = 3.2, p-value = 2×10^−8^, dominance deviation p-value = 10^-5^). It would likely have been missed under the additive model (OR = 1.3, p-value = 9×10^−3^). This example also illustrates the value of applying recessive testing to a cohort enriched for homozygosity. The AF for this variant in G&H is 0.17, which is similar to the AF in non-finnish europeans (NFE) of 0.16 ^73^. The NFE population has low levels of consanguinity, therefore at a sample size of 44,000, one could estimate based on the HWE that the number of homozygous individuals would be 1,126 and the power to perform this recessive test in this population would be 57%. Due to increased autozygosity, the observed number of homozygotes in G&H is 1,408, giving 82% power. Therefore, it is possible that additive tests in larger outbred cohorts have missed this recessive association. An exome-based study of 394,841 UK Biobank individuals and 4,529 phenotypes detected a nominally-significant gene-based additive association between putative Loss-of-Function variants in *MYO16* with the same ICD10 code (OR = 1.1, p-value = 5×10^−4^) ^74^. Furthermore, additive tests with FinnGen (release 6) report nominally significant associations between our lead variant and related phenotypes such as fibroblastic disorders (OR = 1.1, p-value = 10^-3^) and benign neoplasms in the scapula and long bones of the upper limb (OR = 0.7, p-value = 10^-3^) ^30^.

From another study within this cohort, Malawsky et al. (2023) ^75^ demonstrated that increased homozygosity (higher F_ROH_) was associated with multiple common diseases. The primary hypothesis for these associations is that homozygous regions of the genome contained causal variants with recessive effects on these phenotypes ^76^. From the meta-analysis of highly consanguineous cohorts in the study, F_ROH_ was found to be associated with twelve ICD10 subchapters (passing the FDR 5% multiple testing threshold). Five of the twelve (42%) phenotypes significantly associated with F_ROH_ had underlying single-variant recessive associations reported in this study, compared to thirteen of the remaining forty-nine (27%) phenotypes tested in that paper which were not significantly associated with F_ROH_(Fisher’s Exact Test p-value = 0.31). Manually relaxing the phenotype-matching for the twelve phenotypes associated with F_ROH_, we found variants with significant (p<5×10^−8^) recessive associations to three more closely-related phenotypes (Appendix-11), although none passed our more stringent Bonferroni correction threshold. This supports the hypothesis that the association between increased autozygosity and the prevalence of some common diseases is due to underlying genetic variants with recessive effects.

There are several limitations to the project. Firstly, we have not carried out fine-mapping of these recessive associations, because to our knowledge, there are no established methods for fine-mapping which would help disentangle, for example, a recessive hit that is in LD with a strongly additive hit. However, it is worth noting also that LD diminishes with distance between non-additive variants at a rate that is squared of the rate between additive variants ^1^, which should, in theory, make it easier to pinpoint causal recessive variants due to the lower LD. Next, there is limited replication of findings with p-values slightly below the genome-wide significance threshold. This might be because this p-value cutoff is not stringent enough given the number of tests we performed. Also, our power calculations, which were performed on a simple logistic regression model, may have overestimated the power of the model fitted by SAIGE and REGENIE. Furthermore, effect sizes used in the power calculation might be overestimated in the discovery GWAS (winner’s curse), which could also lead to overestimated power. Additionally, differences in the granularity of disease classification and differing methods for phenotype curation are not accounted for in the power calculation. For example, there is no thalassaemia phenotype available for testing in FinnGen (or GERA), and the G&H phenotypes of “thalassaemia” and “other hereditary haemolytic anaemia” were matched to “other anaemia” and “haemolytic anaemia” in FinnGen, which may have contributed to the poorer replication in that cluster of loci compared to the other Bonferroni-significant loci. Lastly, the Finnish cohort and the GERA cohort are composed of very different ancestries from British South Asians, and differences in linkage disequilibrium patterns between populations could be significant. In particular, the relatively poor replication of Bonferroni-significant variants associated with thalassaemia and hereditary anaemias in FinnGen may be because selection for resistance to malaria in South Asia has produced complex LD patterns in that genomic region ^77,78^ which differ from the LD patterns in European-ancestry samples; these are not accounted for in our power calculation. Another limitation is that we only carried out single-variant tests. In future, exome sequencing of the full G&H cohort would allow us to identify rare variants which could be aggregated within a gene to try to boost power to find genes with recessive effects, and reduce the need for fine-mapping.

In conclusion, with our whole-exome and whole-genome imputation sets, we profiled the recessive landscape at single variants in this cohort of 44,000 British South Asians across a broad spectrum of clinical phenotypes, identifying 185 recessive associations. It is likely that many recessive findings remain to be found, and this project provides a sound argument to expand the search to other cohorts and phenotypes.

## Appendices

## Appendix-1: The statistical power needed for recessive analyses

We ran simulations to show that power to detect a recessive effect can be boosted firstly by fitting a recessive rather than an additive model, and secondly by increased homozygosity in the cohort.

We used the genpwr R package ^26^ for the simulations. We applied the following parameters for all calculations: sample size = 44,000 (mimicking G&H), p-value threshold = p<5×10^−8^, model = recessive (or additive), regression = logistic, and assumed there was no gene-environment interaction. The power, allele frequency (AF), case rate and odds ratio (OR) were modified depending on what was being simulated. Since the package does not accept genotype frequencies, but rather, takes the AF as input and assumes the Hardy-Weinberg Equilibrium (HWE) to determine the genotype frequencies, to account for increased autozygosity in G&H, we calculated the frequency at which one would expect to see the number of homozygotes that we would actually see under HWE, denoted below as “AF_autozyg_’’. Specifically, we calculated:

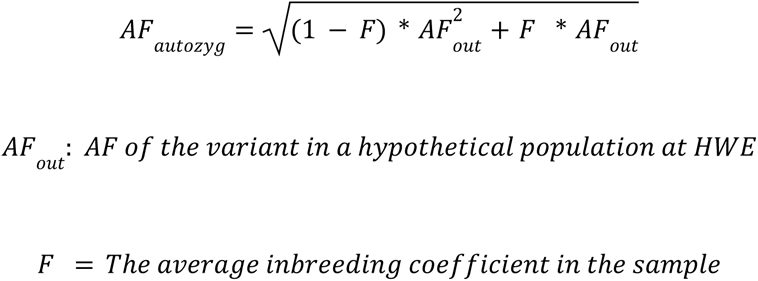

The parameter F (commonly known in the scientific literature as the “inbreeding coefficient”) corresponds to the average relatedness of parents of individuals in the sample with each other. It also corresponds to the average fraction of the genome homozygous in a given sample. ^79^

We assessed if we would have power to detect recessive findings with similar ORs, AFs and case rates as those detected in FinnGen. The FinnGen recessive hits were collated from Table 1 of Heyne et al. (2023) ^8^. We filtered the thirty-one findings to the eighteen findings (Supplementary Table 3) that were validated in release 6 ^11^. The ORs, AFs and case rates were then projected into the power calculations we performed.

Using an OR of 2 and a phenotype case frequency of 1%, we calculated the power of a recessive versus an additive test across the AF spectrum, simulating various levels of average autozygosity in the cohort, ranging from a cohort with no autozygosity (F = 0), to a consanguineous cohort with an average inbreeding coefficient of 10% (which corresponds to the average fraction of individuals homozygous at any position in the genome). In practice, the average fraction of the genome homozygous in individuals from G&H is ∼2.2%. We first see that the additive model had less power for detecting a truly recessive effect than the recessive model, particularly at lower AFs (Figure-S1A,B). Next, we see that power to detect this recessive effect increased with the average level of autozygosity, and for this set of parameters, this was most noticeable around the AF 0.2 - 0.5 range (Figure-S1A). This sample size and levels of consanguinity simulated were not powered to detect an effect size of OR = 2 in rare variants, therefore we increased the OR to 5 and reran the simulations for the rare and low frequency spectrum, and we still saw that power is higher with increasing F (Figure-S1B).

**Figure S1.**
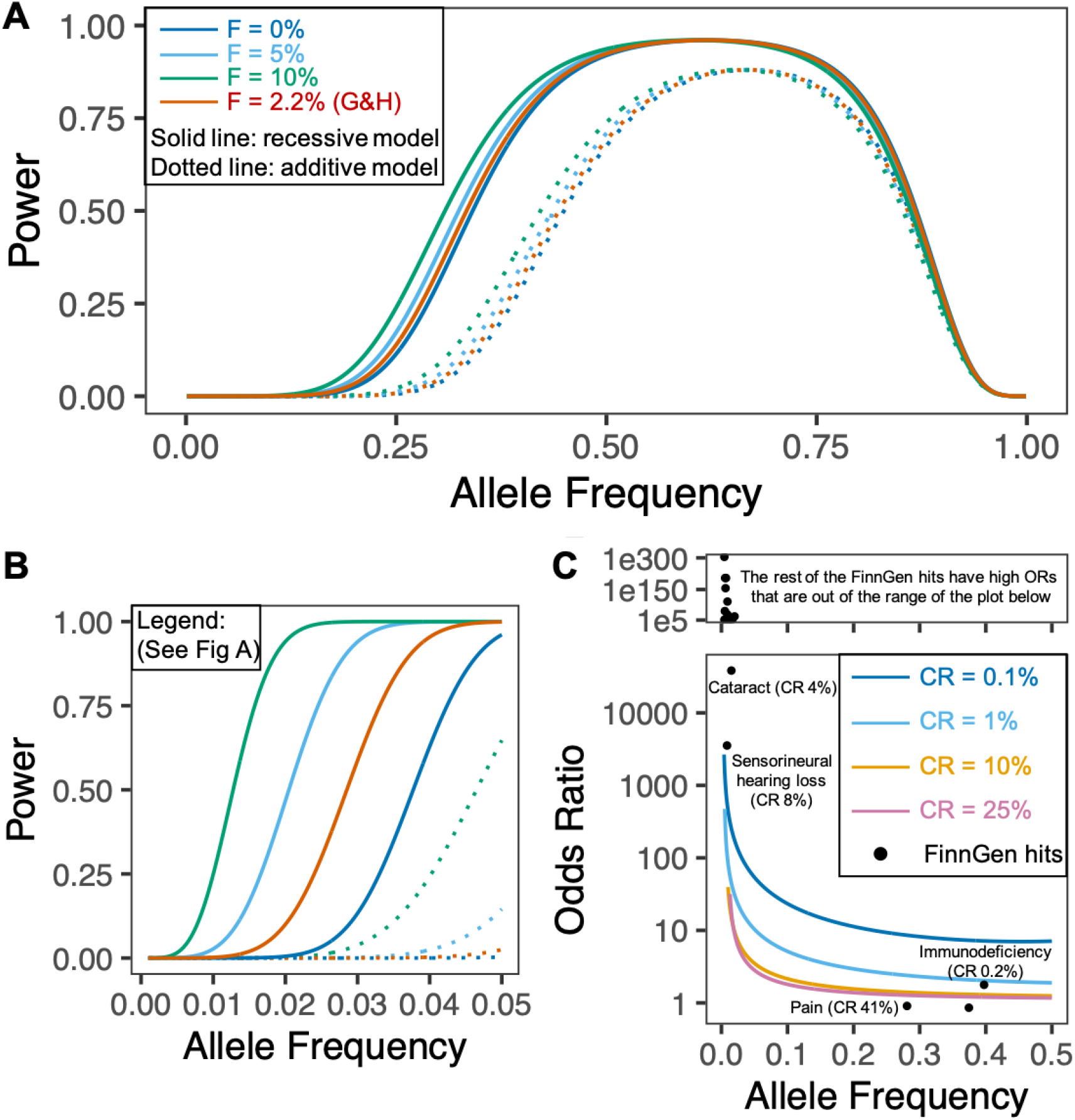
Simulations to evaluate power to detect a recessive effect in G&H when using a recessive versus additive model. A: Calculating power to detect an association at p<5×10^−8^ for a range of allele frequencies and values for average inbreeding coefficient (F). Sample size = 44,000, OR = 2, CR = 1%. B: Calculating power for a range of rarer allele frequencies and values for F. Parameters as per A, except OR = 5. C: Calculating the minimum odds ratio G&H would be powered to detect at 80% power, and F = 2.2%. ”CR” : Case rate. Recessive hits reported in FinnGen ^8^ are indicated on the plot, showing their OR and AF estimated in FinnGen.

We next wanted to evaluate the minimum OR we would be powered (80% power) to detect in G&H. To do that we simulated varying case rates from 0.1% to 25%. We projected the recessive findings reported in FinnGen by Heyne et al. (2023) onto our simulations, and found that the majority of the findings had ORs higher than the minimum that we are powered to detect at their corresponding case rates and AFs (Figure-S1C). This suggested that G&H will be well powered to detect very large ORs at rare variants i.e. effectively Mendelian associations, and smaller recessive effects at common variants with common traits.

## Appendix-2: Flowchart of methods

**Figure S2.**
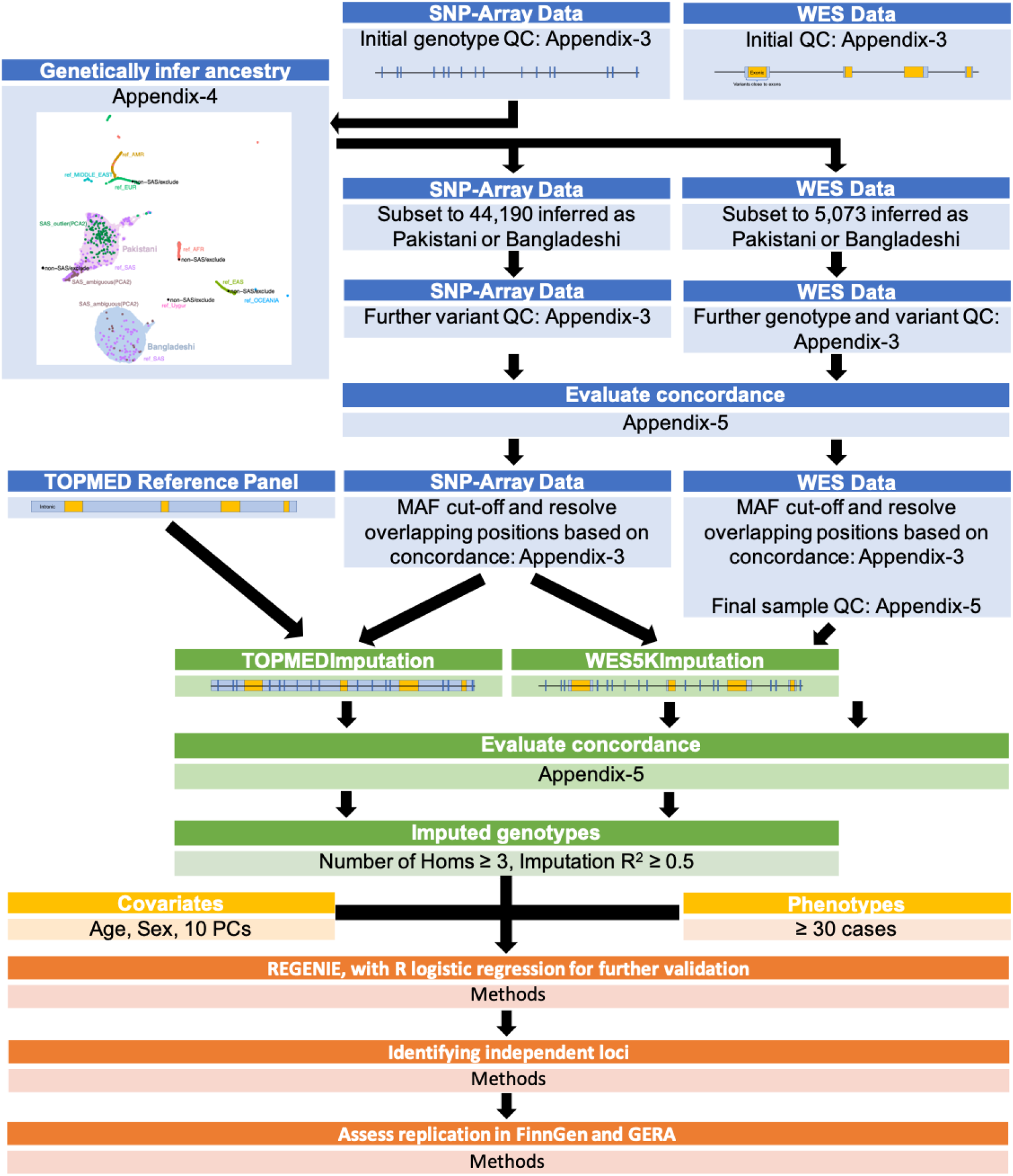
A flowchart to visualise the methods of this project. This is intended to serve as a directory to relevant sections.

## Appendix-3: QC of genetic data

**Table S1.** QC steps for the SNP-Array and WES Data.

|  | SNP-Array Data | WES Data |
| --- | --- | --- |
| **Initial QC** | **Initial genotype QC**<br>44,396 individuals<br>Variant QC in GenomeStudio<br>Sample QC (exclude samples with low call rate or failed sex checks) | 5,236 individuals after light sample QC (exclude samples with low coverage or sex discrepancies) |
| **Genetically infer ancestry** | 44,190 individuals assigned as Pakistani or Bangladeshi | 5,073 individuals with array data and assigned as Pakistani or Bangladeshi |
| **Genotype QC** |  | Set genotype as missing when: GQ <20, binomAD <10<sup>-2</sup>, DP ≤7 |
| **Variant QC** | 534,806 autosomal SNPs | 2,245,547 autosomal SNPs and indels, after excluding:<br>For SNPs: "QD < 2.0 \|\| FS > 30 \|\| MQ < 40.0 \|\| MQRankSum < -12.5 \|\| ReadPosRankSum < -8.0"<br>For indels: "QD < 2.0 \|\| FS > 30 \|\| ReadPosRankSum < -20.0" |
|  | 534,531 variants with a call rate ≥99% | 1,645,161 variants with a post-genotype QC call rate ≥70% |
|  | 533,362 variants with HWE p ≥10<sup>-6</sup> in the Bangladeshi subgroup |  |
|  | 533,166 biallelic variants |  |
| *Evaluate Concordance* |  |  |
| **MAF cut-offs** | Keep 469,678 variants with MAF >0.1% | Remove singletons |
| **Resolve Overlapping Positions** | Exclude 25 common palindromic variants with MAF >40% | Keep, as no problem of allele-switching |
|  | Exclude 515 variants with overlapping positions but unmatched alleles | Keep, as some are indels that have been genotyped incorrectly on the array |
|  | Keep 38,248 variants with matched alleles and MAF >0.1% | Exclude, as the array has a higher overall call rate<br>(Note: rare variants with MAF ≤0.1% that overlapped with the array are retained) |
| **Final Sample QC** |  | 4,982 individuals after excluding:<br>Pre-genotype QC call rate <4SD from mean, Post-genotype QC call rate <2SD from mean, Post-genotype QC NRD >4SD from mean |

## Appendix-4: Genetic inference of ancestry

We genetically inferred ancestry for the 44,396 individuals by merging the SNP-array data with reference cohorts.

From the SNP-array had been through initial QC, we filtered to variants that were autosomal, common (defined as a minor allele frequency, MAF>0.01), had a call rate of ≥99%, and passed the HWE exact test (p>10^-6^) in self-declared Bangladeshi individuals (Table S1). This filtered dataset was then merged with reference sequences of 3,433 individuals from the 1000 Genome Project (1000G) ^80^ and Central and South Asian individuals from the Human Genome Diversity Project (HGDP) ^81^. We excluded palindromic variants, and variants with significant AF differences between G&H and 676 reference South Asians curated from 1000G and HGDP (since these represented likely genotyping errors). We defined variants with significant AF differences in the following manner: Firstly, we calculated the residuals from the linear regression between the AFs in both datasets. Next, we binned the variants into bins by frequency (in intervals of 0.01), and selected variants for which the residual was >5 standard deviations (5SD) away from the mean of the residuals in that frequency bin. (This choice seemed reasonable after testing various SD thresholds, Figure-S3A). Lastly, we performed Fisher’s exact tests to compare the genotype counts between the G&H data and the 676 reference South Asians at the variants selected above and excluded those with a p<10^-5^. (Again, we tested p-value thresholds ranging from 0.05 to a multiple testing correction of <0.05/349,632 variants, and felt that 10^-5^ was reasonable, Figure-S3B). The various thresholds and the distribution of the outlier variants excluded are graphically represented in Figure-S3C-D.

**Figure S3.**
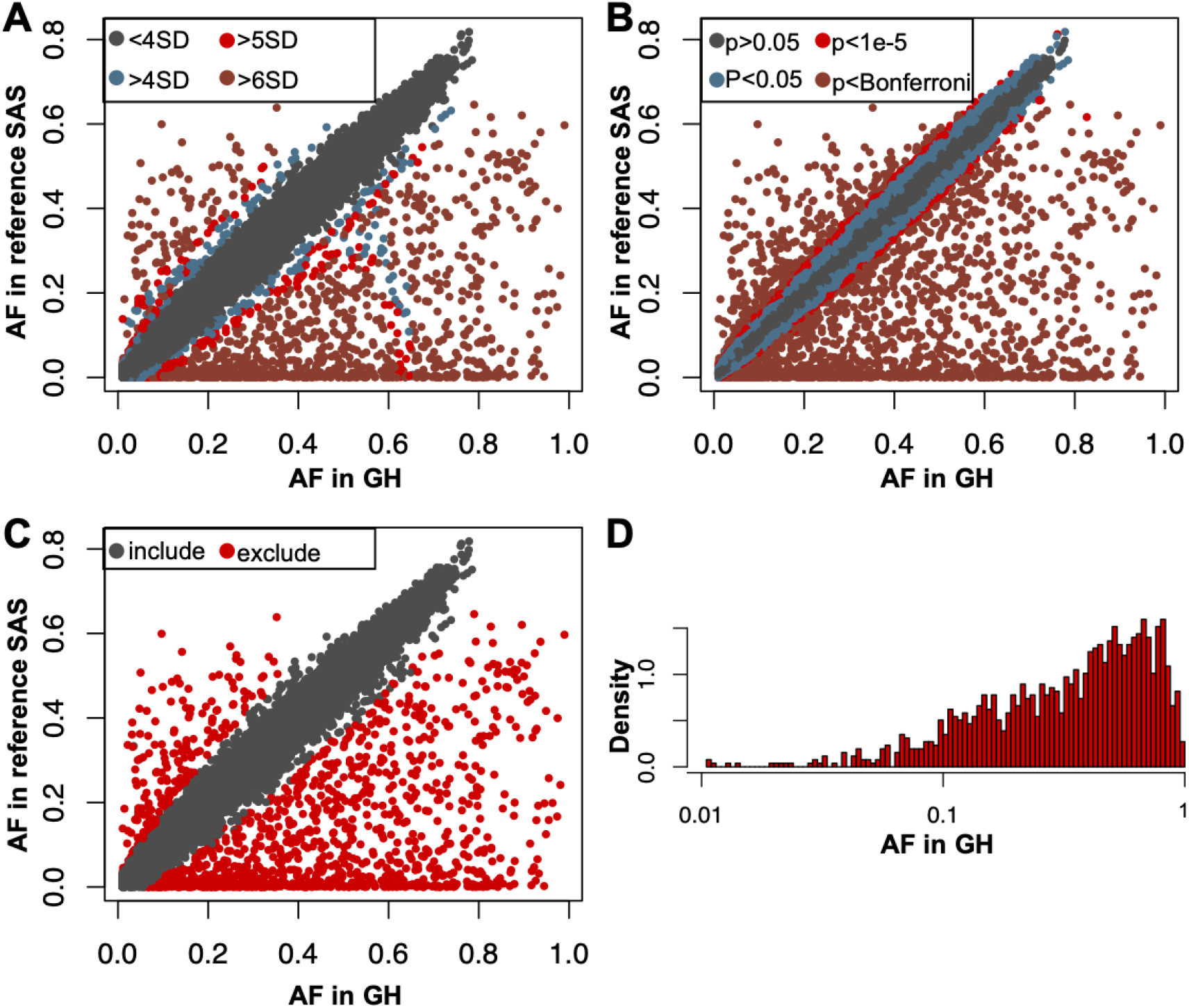
Excluding variants with significantly different AF in Genes and Health (GH) and 676 South Asian samples from 1000 Genomes and HGDP (reference SAS). Plotting the AFs in both cohorts for each variant, colour-coding represents in A: the different standard deviation (SD) thresholds applied to the mean of the residuals in 0.01 frequency bins, in B: the different p-value thresholds for Fisher’s exact tests on the genotypes, and in C: the variants that were included and excluded from the merged panel of variants. D: The AF distribution of the variants excluded.

After merging of G&H with the reference samples, LD pruning was performed (window size 1000 kilobases (kb), step size 50, LD R^2^ 0.1) with PLINK1.9 and long LD regions were excluded. ^82^

Principal component analysis (PCA) was performed with PLINK1.9 on the reference individuals, then the G&H individuals were projected into the reference PC space (PCA1). We calculated uniform manifold approximation and projection (UMAP) coordinates (umap R package) ^83^. We found that the UMAP with 7 PCs was optimal to separate the reference individuals into superpopulations. 44,320 out of 44,396 G&H individuals were inferred to be South Asian at this stage (Figure-S4A), and carried forward for the downstream analysis.

The PropIBD algorithm in KING ^84^ was run to estimate pairwise relationships up to fourth degree within G&H, and we removed a minimal set of 14,727 individuals who had at least one relative (3^rd^ degree and closer) in the dataset, leaving 29,668 unrelated individuals.

We performed a second PCA (PCA2) on the unrelated G&H individuals, projecting the related G&H individuals who we had inferred to be South Asian into the PC space. The UMAP with 4 PCs identified distinct clusters that corresponded well to self-declared Bangladeshi/Pakistani ancestry, and this was used to genetically classify individuals as genetically Bangladeshi or Pakistani (Figure-S4B).

**Figure S4.**
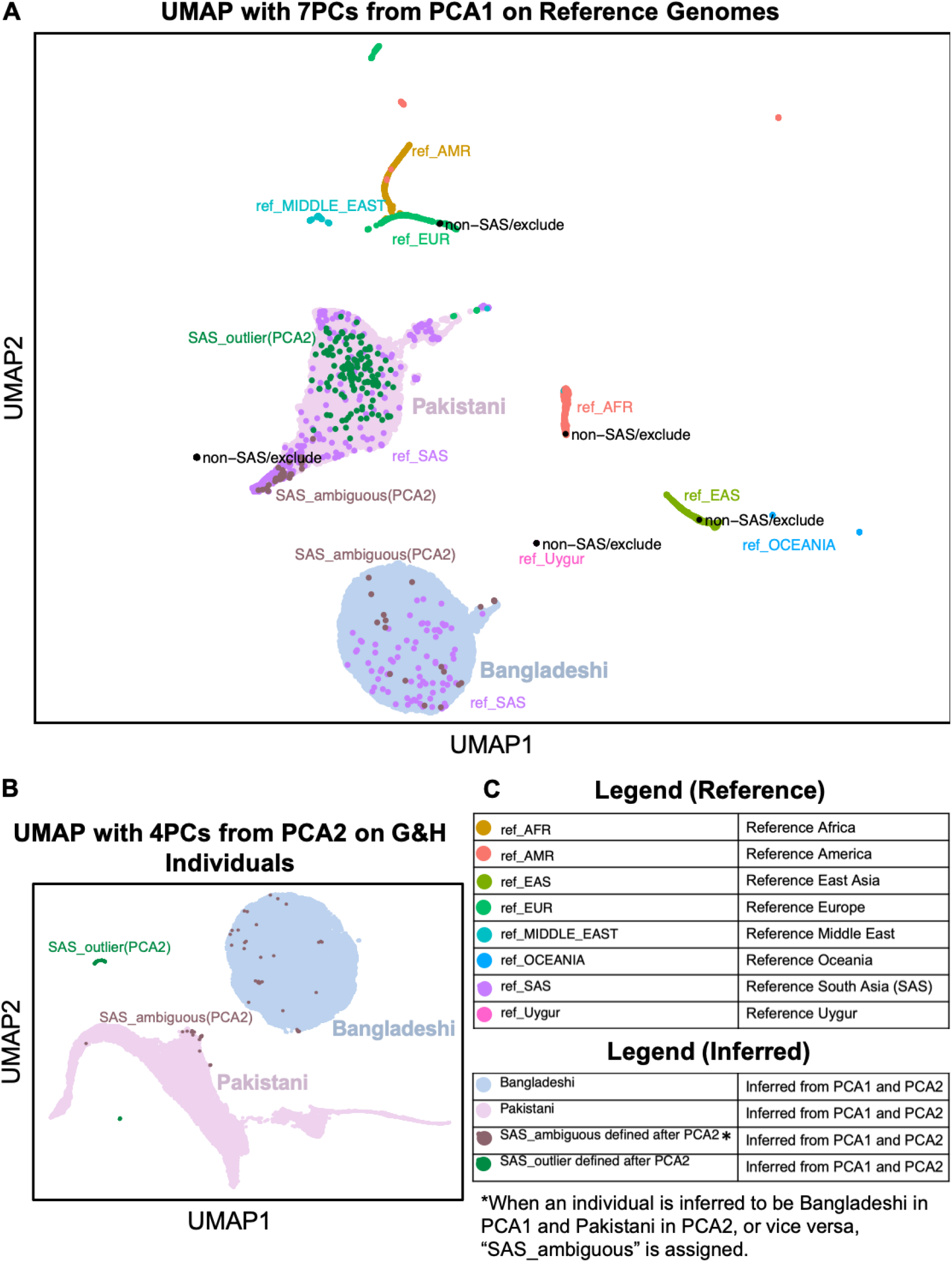
Genetic inference of ancestry using the G&H SNP-array data. A: UMAP with 7 PCs from the PCA of reference individuals (PCA1), with G&H individuals projected into the PC space. B: UMAP with 4 PCs from the PCA of unrelated G&H individuals (PCA2), with related G&H individuals projected into the PC space. C: Legends for both UMAPs.

## Appendix-5: Assessing genotyping and imputation accuracies with concordance analyses

Before merging the SNP-array and WES data to build the reference panel, we evaluated concordance of genotypes between them at overlapping sites.

Furthermore, after imputation, we assessed imputation accuracy by comparing imputed genotypes to sequenced genotypes.

### Evaluating concordance

Concordance was primarily evaluated by the non-reference discordance rate (NRD) calculated using the following formula:

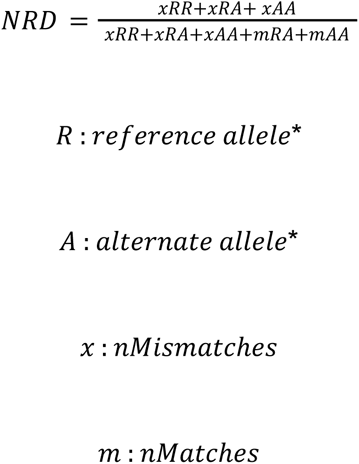

*Note that the ‘truth’ was defined by the array genotype when comparing concordance between array and WES data, and defined by the sequenced genotype when comparing concordance between sequenced and imputed data. For example, “xRR” would mean that the truth dataset had a homozygous

reference genotype while the other dataset did not, and “mRA” means that both datasets had a heterozygous genotype.

When examining NRD stratified by allele frequency, the NRD was modified to calculate a “minor allele discordance” (MAD) rate:

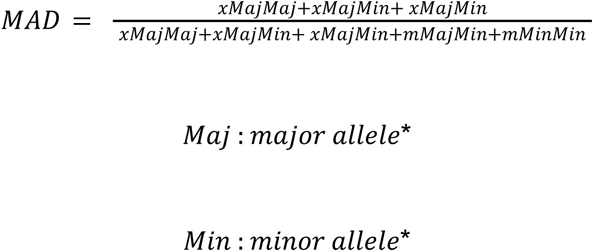

Where relevant, Pearson correlation R^2^ between genotypes / dosages were calculated as a secondary measure of concordance.

### Concordance between the SNP-array and the WES data

QC on both the SNP-Array and WES datasets improved the overall NRD from 5.02% to 0.40%, with the WES GQ filter making the biggest difference (Table S2). We anticipated that the genotyping accuracy would be low for rare variants in the SNP-array, but surprisingly the array data continued to demonstrate good MADs of ∼0.7-0.9% with the WES for variants with minor allele counts (MACs) on the array of 3-6 (equivalent to MAF 0.03-0.06%, Figure S5). We decided to use variants with MAF>0.1% from the SNP array for the imputation backbone.

Since minimac3 (Which is used to build the reference panel for imputation with minimac4) does not tolerate missingness in the imputation reference panel, we needed to use the WES data without any genotype-level QC to minimise missingness. To select for variants with a majority of high-quality genotypes, we retained those that had <30% missing genotypes after applying the genotype-level QC. Using the raw genotypes at those sites increased the NRD to 1.26% (Table S2). Note that this value is inflated by the discordance at sites with MAC 0-2 (Figure S5), but variants this rare were filtered out of the SNP-array when building the imputation backbone, and instead retained in the WES data, since we assume these are likely to be more accurate than the array genotypes given the difficulties of genotyping uber-rare variants on arrays ^85^.

**Table S2.** Overall non-reference discordance (NRD) between the array and WES data with different QC filters applied to the SNP-array data and the WES data. Row names describe stages of SNP-array filtering, column names describe stages of WES filtering. “Initial genotype QC” is described in the first row of Table S1.

| | Raw Genotypes<br>(GTs) | GQ $\geq 20$ | Call rate $\geq 70\%$<br>after:<br>GQ $\geq 20$<br>binomAD $\geq 10^{-2}$<br>DP $> 7$ | With raw GT at<br>sites with call rate<br>$\geq 70\%$ after:<br>GQ $\geq 20$<br>binomAD $\geq 10^{-2}$<br>DP $> 7$ |
| --- | --- | --- | --- | --- |
| After initial<br>genotype QC | 5.018%<br>(46,778 SNPs) |  |  |  |
| Call rate $\geq 99\%$<br>HWE pval $\geq 10^{-6}$ | 5.000%<br>(46,656 SNPs) | 0.500 %<br>(46,401 SNPs) | 0.398%<br>(38,249 SNPs) | 1.258 %<br>(38,249 SNPs) |

**Figure S5.**
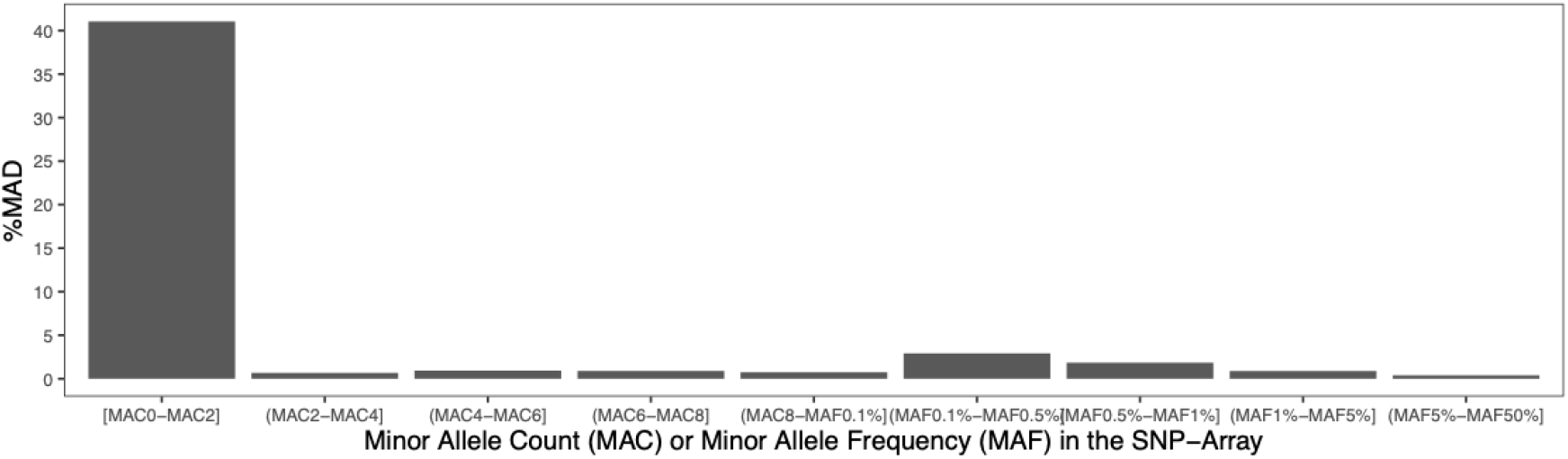
The Minor Allele Discordance (MAD) between the SNP-Array and the WES at overlapping variants, stratified by the frequency on the array. Array filters: After initial genotype QC + Call rate ≥99% + HWE pval ≥10^-6^. WES filters: Call rate ≥70% after GQ ≥20 + binomAD ≥10^-2^ + DP>7. Here, the array genotypes were treated as “truth”.

Based on the concordance analysis, we decided to include only array variants with MAF>0.1% in the imputation backbone. Overlapping positions with the WES were resolved as follows:

- For SNPs at overlapping positions with matched alleles and with MAF>0.1%, we retained these in the GSA data but removed them from the WES.
- We excluded overlapping common palindromic variants (MAF>0.4) where the strand could not be confidently determined.
- We excluded overlapping positions with unmatched alleles (including all indels).

This resulted in 469,678 variants, which were then phased with EAGLE2 (Kpbwt=20,000) ^20^.

Based on the concordance of the SNP-array and WES, 91 samples that had high missingness and/or high non-reference discordance (NRD) values were excluded (Figure-S6B, Figure-S6C), leaving 4,982 samples. Specifically, we excluded individuals who had any of the following:

- a raw WES call rate <4SD from the mean raw WES call rate,
- a post-genotype QC WES call rate <2SD from the mean post-genotype QC WES call rate,
- an NRD based on the post-genotype QC WES and post-QC SNP-array data of >4SD from the mean

To build the reference panel, we filtered the WES data to a subset of high-quality variants that had ≥70% call rate after genotype-level QC, and removed singletons. Since Minimac3 does not cope with missing genotypes, we used the genotypes at those sites from the raw data (i.e. pre-genotype QC), and replaced missing genotypes (∼0.02% of the total genotypes, Figure-S7A) with 0/0 if the reference allele was the major allele and 1/1 otherwise. The distribution of raw per-variant missingness at sites that pass a post-genotype QC call rate ≥70% is shown in Figure-S7B, with most variants having a raw missingness of <1%.

**Figure S6.**
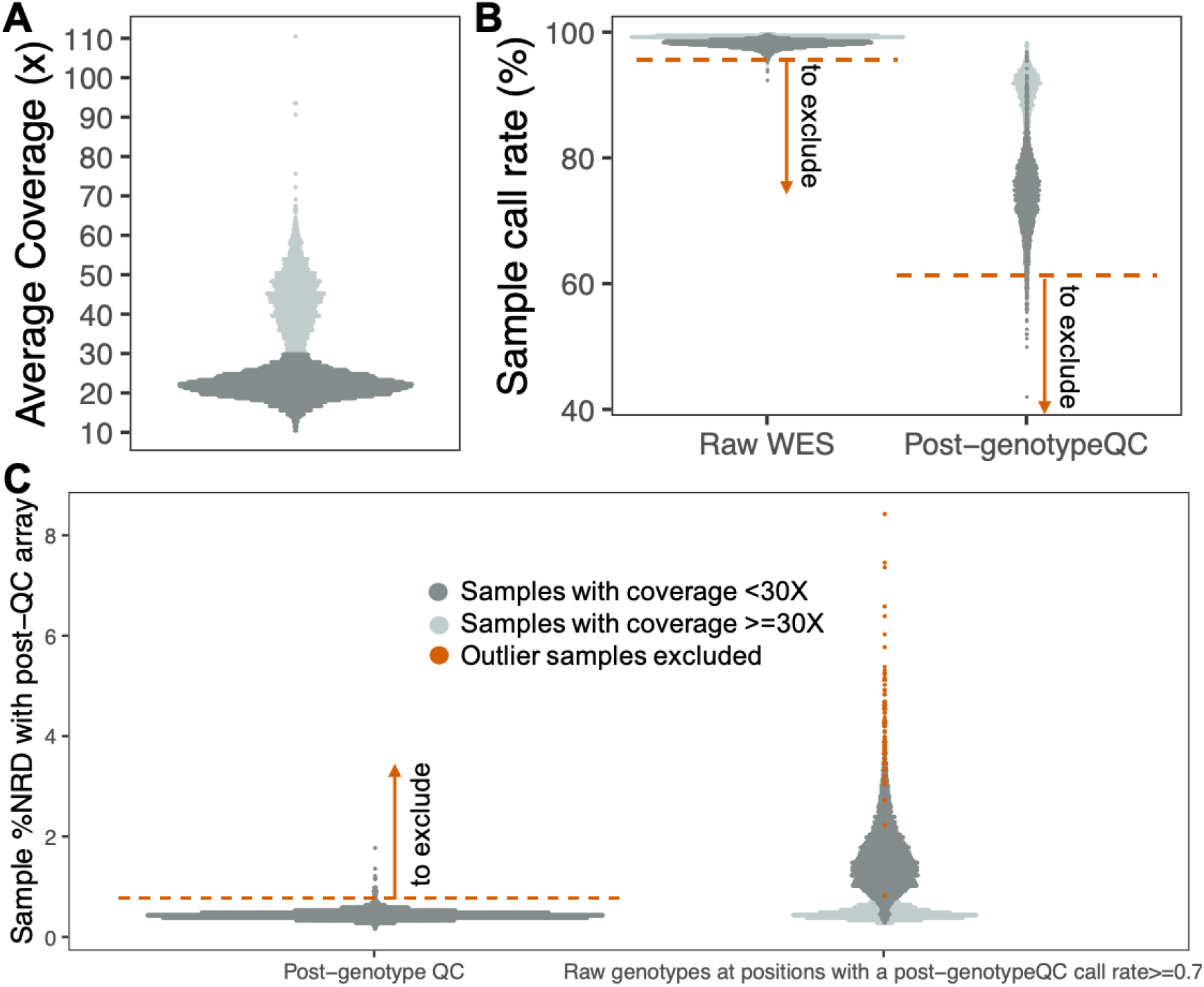
Per-sample coverage, call rate, and non-reference discordance (NRD) before and after QC of WES data. A: Distribution of average on-target coverage across samples. The bimodality observed is because different batches of samples were sequenced to either ∼20X or ∼40X. B: The distribution of WES sample call rates pre-genotype QC (left) and post-genotype QC (right). C: The distribution of NRDs across samples in the post-genotype QC WES. In B and C, the bimodality is again likely due to the different sequencing coverage.

**Figure S7.**
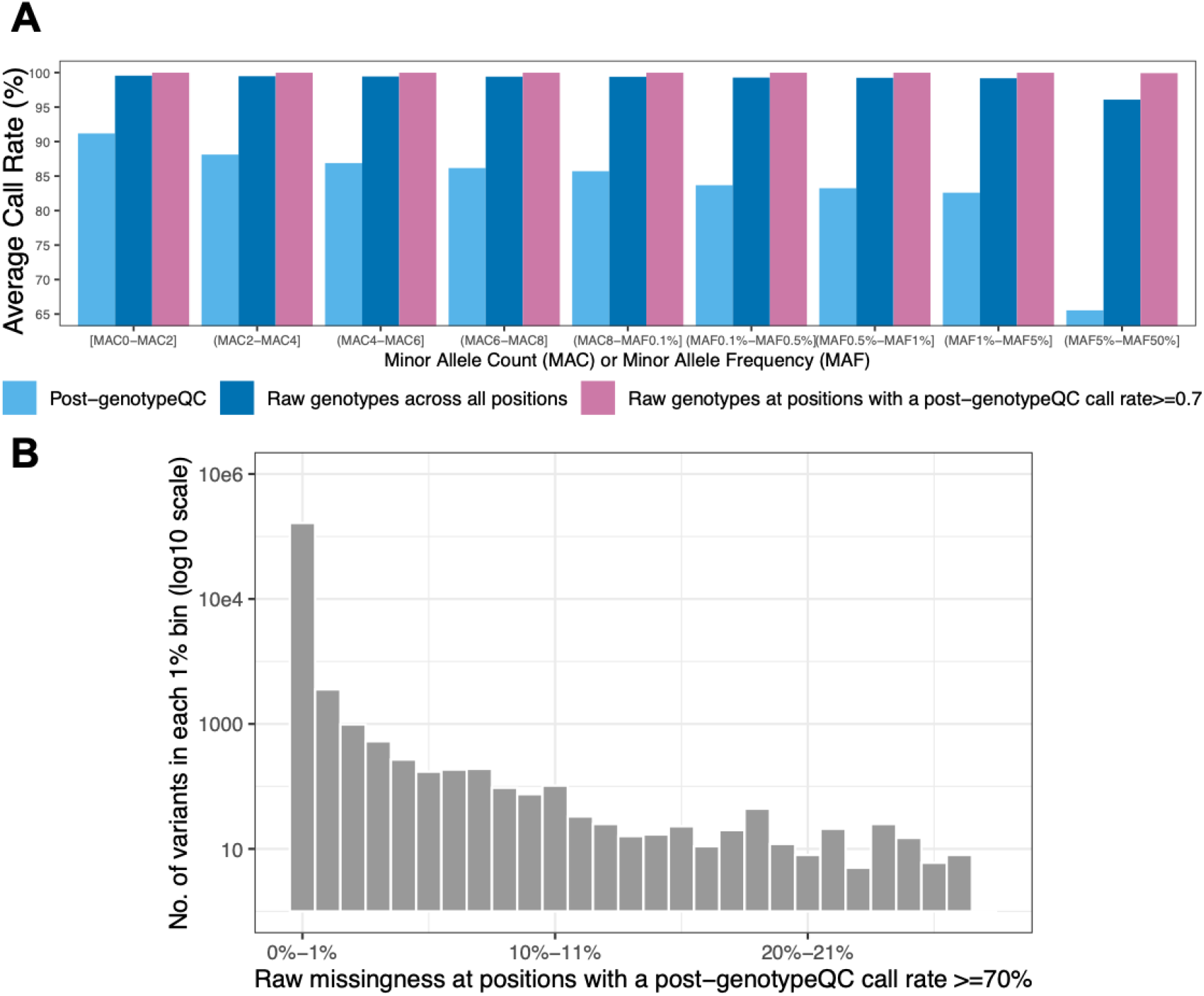
Distribution of call rates pre and post QC in the G&H WES data. A: Average call rate stratified across MAF bins. As the missingness is low in the raw WES, when filtering to positions with a post-genotypeQC call rate ≥70%, the call rate of the raw genotypes is >99% across the frequency spectrum. B: Distribution of per-variant raw missingness at positions with a post-genotype QC call rate ≥70%. The y-axis has been transformed to a log 10 scale as an overwhelmingly large number of variants have a raw missingness of 0-1%.

Variants with MAF>0.1% that were also present on the GSA array and had been included in the the imputation backbone were removed from the reference panel, since they tended to have higher call rates in the array data. The cleaned WES and cleaned SNP-array data from the 4,982 samples were then merged to form the reference panel and phased with EAGLE2 (Kpbwt=20,000). The reference panel consisted of 1,385,942 variants.

### Evaluating imputation accuracy

After imputation, to assess the imputation accuracy, the imputed genotypes for the 4,982 individuals with WES data were compared to their sequenced genotypes. Genotype concordance was evaluated between the SNP-array and the WES data at overlapping sites, and between the sequenced and imputed variants.

Ten trials of imputation were performed using the WES5K panel, with each trial leaving 10% of the WES samples out of the reference panel against which we then evaluated concordance between the sequenced and imputed genotypes.

The total number of variants in each imputed dataset ranged from 1,325,855-1,327,697 across the 10 trials. Although the genotyped backbone is imputed by Minimac4 as well, to evaluate imputation accuracy purely at positions with no prior information, we excluded backbone SNPs resulting in 855,697-857,537 variants to compare in each trial. The overall NRD of the imputed genotypes compared to the sequenced genotypes ranged from 7.44-7.69% across 10 trials (Figure-S8A).

The confidence of the imputation is quantified by Minimac4’s imputed R^2^. By convention, a minimum cutoff of ≥0.3 is applied to QC imputed data, which should sufficiently filter out poorly-imputed variants based on the distribution of imputed R^2^ scores (Figure-S8B). However the overall NRD was not found to improve at this cutoff (Figure-S8A) (probably because we are including so many rare variants) so we applied a more stringent cutoff of ≥0.5. As expected, the rarer MAF bins had more variants with a lower imputed R^2^ (Figure-S8C).

MAD also increases with decreasing allele frequencies, as expected. Many of these extremely rare variants will not be submitted for association testing as they are both poorly imputed and not powered enough for recessive tests.

After applying imputed R^2^ ≥0.5 and N_Hom_ ≥3 to prepare for association tests, and including the positions of the SNP-array backbone (which will be included in association tests), the overall NRD improved to 1.78-1.84% across the trials (Figure S9).

**Figure S8.**
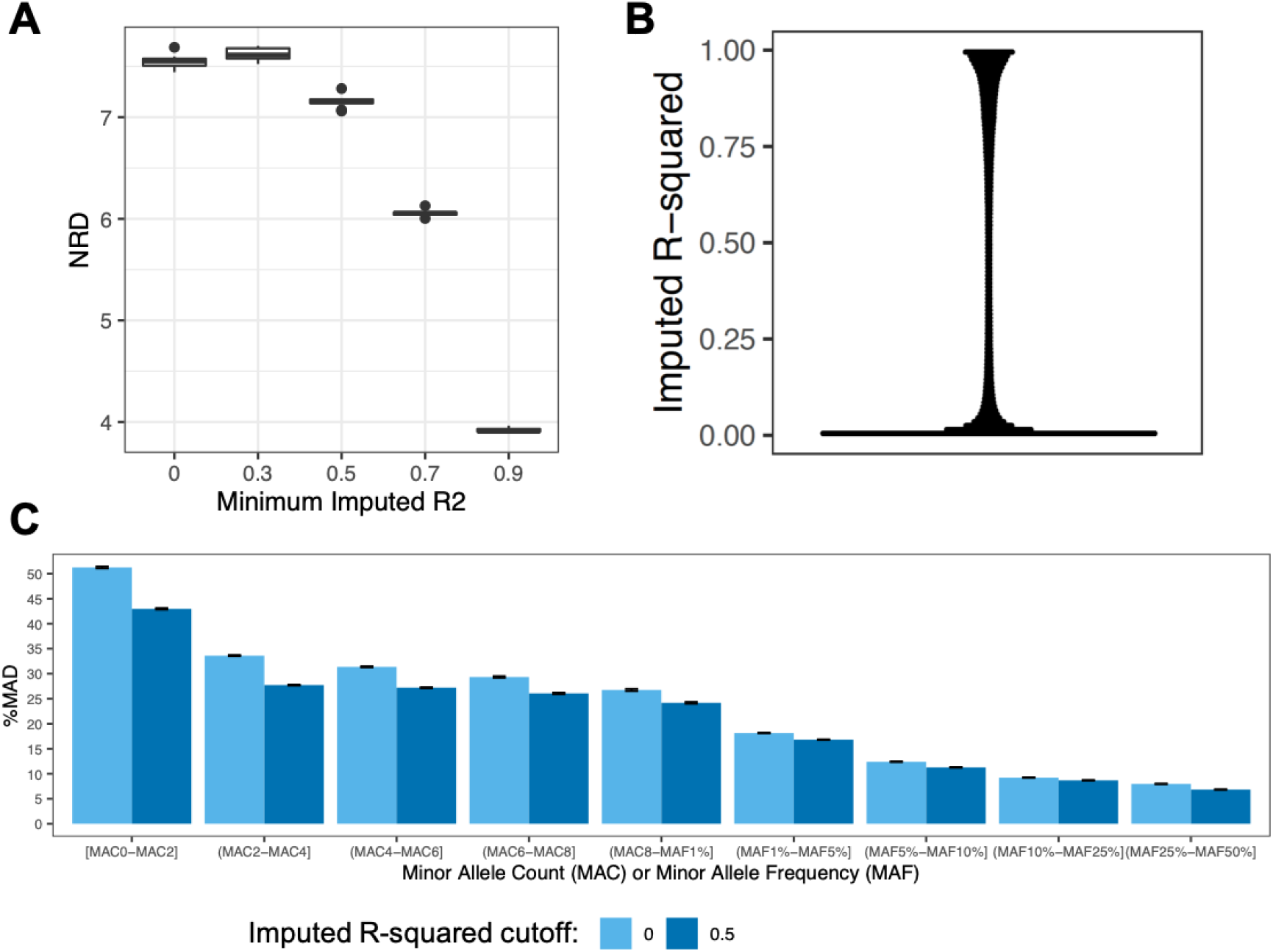
Results of the leave-10%-out trials. A: The distribution of NRD(%) across 10 trials when different minimum imputed R^2^ cutoffs are applied. B: The distribution of imputed R^2^ scores for a representative trial. C: The MAD before and after applying an imputed R^2^ ≥0.5 cut-off, stratified by MAF. (Error bars are the standard errors in %MAD across the 10 trials.)

The TOPMEDimputation genotypes of the 4,982 samples with WES were compared to their sequenced genotypes. The same cutoffs (imputed R^2^ ≥0.5 and N_Hom_ ≥3) were applied to result in 10,045,406 variants. Of these variants, 523,018 variants overlapped with the WES and at these, the overall NRD was 1.19%.

We then considered NRD at variants stratified by the number of homozygotes as defined in the WES5Kimputation, the rationale being that the WES should be considered “truth” in this case, and that the power of the recessive association testing depends on the number of homozygotes rather than directly on the allele frequency. First, we observed that the within-cohort WES5K reference panel allowed for more variants to be imputed at the chosen level of accuracy, especially at lower MAFs (Table S3). Second, for N_Hom_ ≥9 up to a homozygous frequency of 5%, the MAD is lower for the WES5KImputation compared to the TOPMEDImputation, but the opposite is true for the lowest and highest MAF bins. Thirdly, the lower overall NRD of the TOPMEDimputation (1.19% versus ∼1.7% for the WES5K Panel) is driven by the improved accuracy of common variant imputation with the TOPMED reference panel. (Figure S9) As both imputation sets carried their own strengths and weaknesses across the frequency spectrum, it was decided to bring both sets forward to association testing.

**Figure S9.**
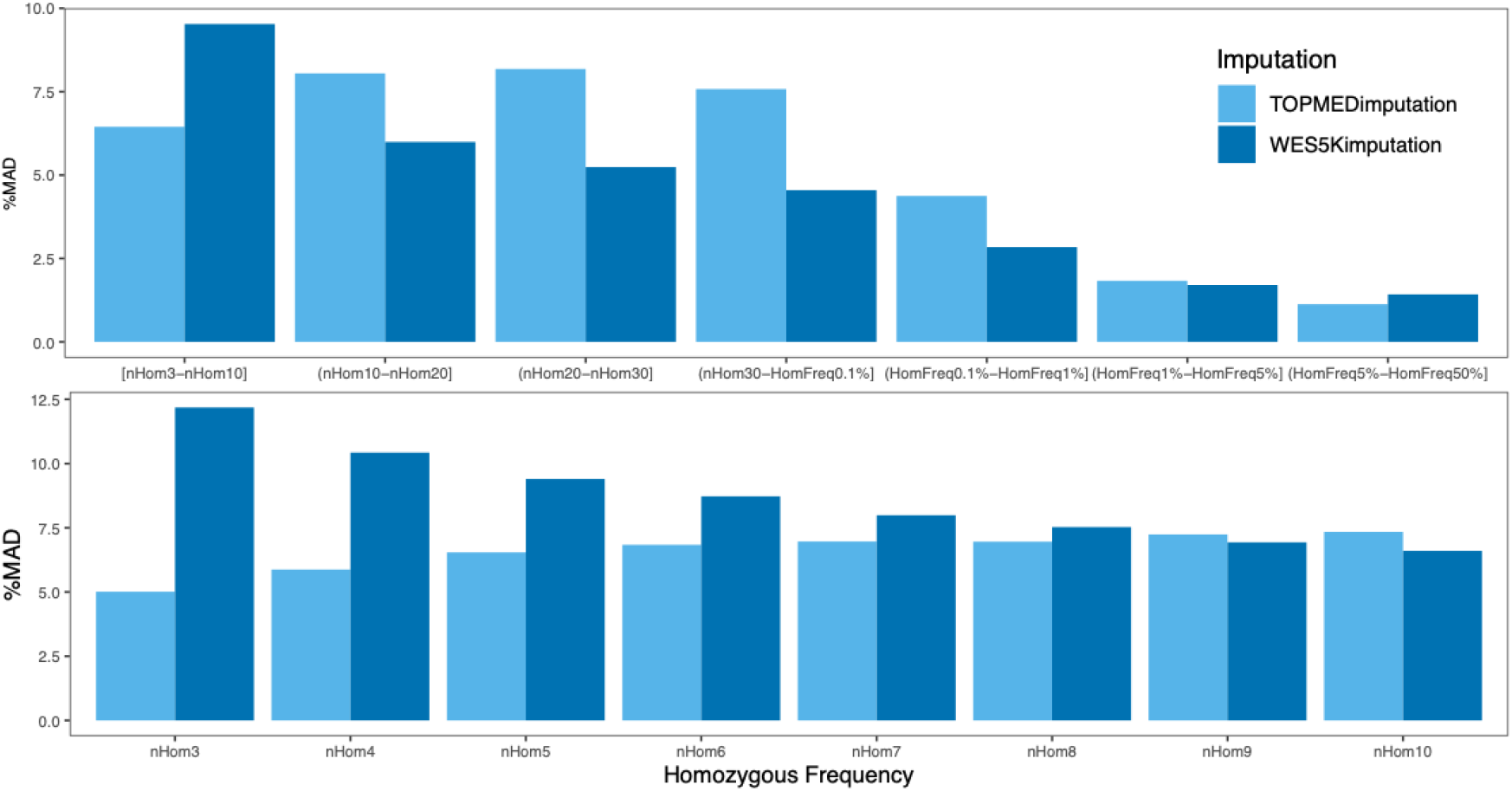
Minor allele discordance (MAD) of the TOPMEDimputation compared to the WES, and the mean MAD across ten leave-10%-out trials compared to the WES (represented as the WES5Kimputation for simplicity), stratified by the number or frequency of homozygous genotypes in the WES data. The variant counts contributing to each bin is tabulated below.

**Table S3.**
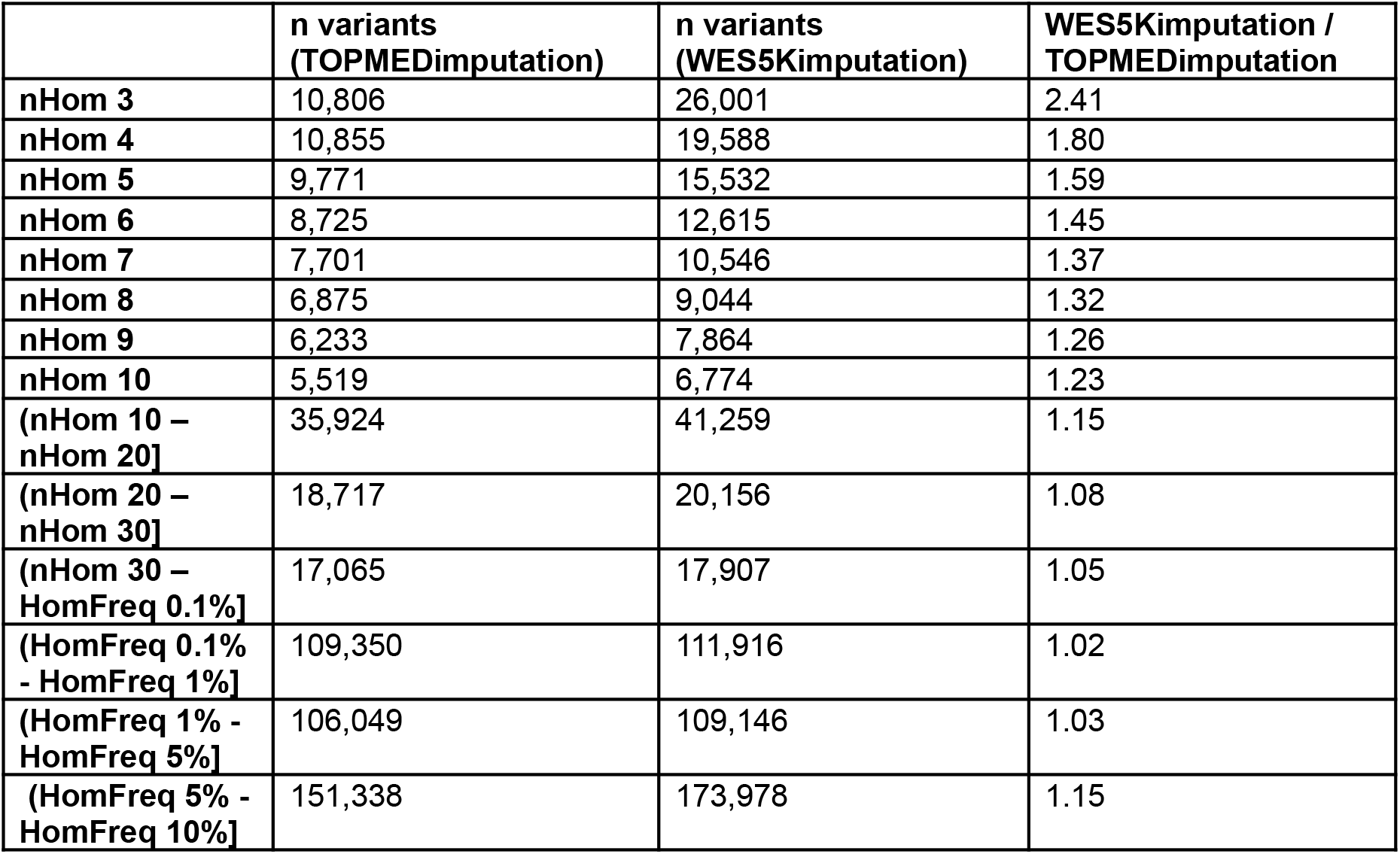
Number of imputed variants (that can be compared to WES positions for concordance analyses) binned by the number or frequency of homozygous genotypes in the WES data.

## Appendix-6: On multiple testing and the independence of phenotypes

The 898 phenotypes tested are not completely independent of each other, and there may be significant correlation between phenotypes, particularly as some diseases are repeated in the custom list and the ICD10 codes. We sought to calculate Pearson correlation R^2^ between phenotype pairs to quantify the degree of correlation in our phenotypic data. This serves two purposes - to evaluate if phenotypes we would expect to be highly correlated indeed have a high R^2^ (meaning that they have been curated correctly), and secondly, to try to eliminate highly-correlated phenotypes to reduce the multiple-testing burden.

Of the 402,753 pairs of phenotypes generated, 153 pairs had a correlation R^2^ of ≥ 0.5. Manually inspecting these pairs, the majority were between conditions one would expect to be highly correlated; for example, the ICD10 encoding for sarcoidosis and for multiple sclerosis fully correlated (r^2^ = 1) with the respective custom encodings for these conditions. There were also correlations between biologically similar phenotypes, such as pulmonary heart disease and pulmonary hypertension (r^2^ = 0.89), and correlations between pairs for which one trait was a subset of the other, such as acute pancreatitis and pancreatitis (r^2^ = 0.90). By reviewing these highly-correlated pairs, we estimated that only about 80-90 phenotypes could be excluded due to being highly correlated, as the rest of the pairs had differences in their definitions that warranted the inclusion of both phenotypes.

We therefore tested all 898 phenotypes available, and to be stringent, the Bonferonni cutoff we used accounted for all tests as if they were independent.

The number of phenotypes, individuals, and variants tested is summarised in Table S4. A small minority of tests failed on REGENIE; a total of 9,197,933,046 tests produced a p-value, out of a possible 9,374,352,233 tests (98% success rate).

**Table S4.** The number of phenotypes, individuals, and variants being tested. The number of variants tested varied with the number of individuals tested, as different subsets of individuals would result in different numbers of variants having at least three homozygotes.

| **Phenotypes** | **n**<br>**phenotypes** | **n**<br>**individuals** | **n variants**<br>**(WES5KImputation)** | **n variants**<br>**(TOPMEDImputation)** |
| --- | --- | --- | --- | --- |
| Custom<br>phenotypes<br>(In both sexes) | 199 | 44,186 | 605,263 | 10,045,406 |
| Custom<br>phenotypes<br>(Female only) | 15 | 24,387 | 553,716 | 9,215,675 |
| Custom<br>phenotypes<br>(Male only) | 5 | 19,799 | 529,480 | 8,855,245 |
| ICD10 codes<br>(In both sexes) | 565 | 42,027 | 599,758 | 9,948,708 |
| ICD10 codes<br>(Female only) | 99 | 23,710 | 550,988 | 9,171,960 |
| ICD10 codes<br>(Male only) | 15 | 18,317 | 522,117 | 8,741,181 |

## Appendix-7: On covariates included in the association testing

The covariates included were age (at year of phenotype curation, 2022), sex, age^2^, age x sex, age^2^ x sex and the first ten principal components (PCs) from the principal component analysis on unrelated G&H individuals described above.

We controlled for ten genetic PCs derived from common variants. These top ten PCs explained more than 85% of the variance explained by the top 50 PCs (Figure-S10A). ^86^ suggested that the common variant PCs may not sufficiently control for the population structure captured within rare variants, and as this study tested rare variants as well, we considered the possibility of controlling for PCs derived from rare variants. We filtered the SNP-array to variants with a minor allele count of 3 to a minor allele frequency of 1%, and completed the QC and PC calculation in the same way as we did for the common variant PCs (described in Appendix-3). Most of the top 10 rare variant PCs were strongly correlated with at least one of the common variant PCs (Figure-S10B). We therefore decided not include rare variant PCs as covariates.

**Figure S10.**
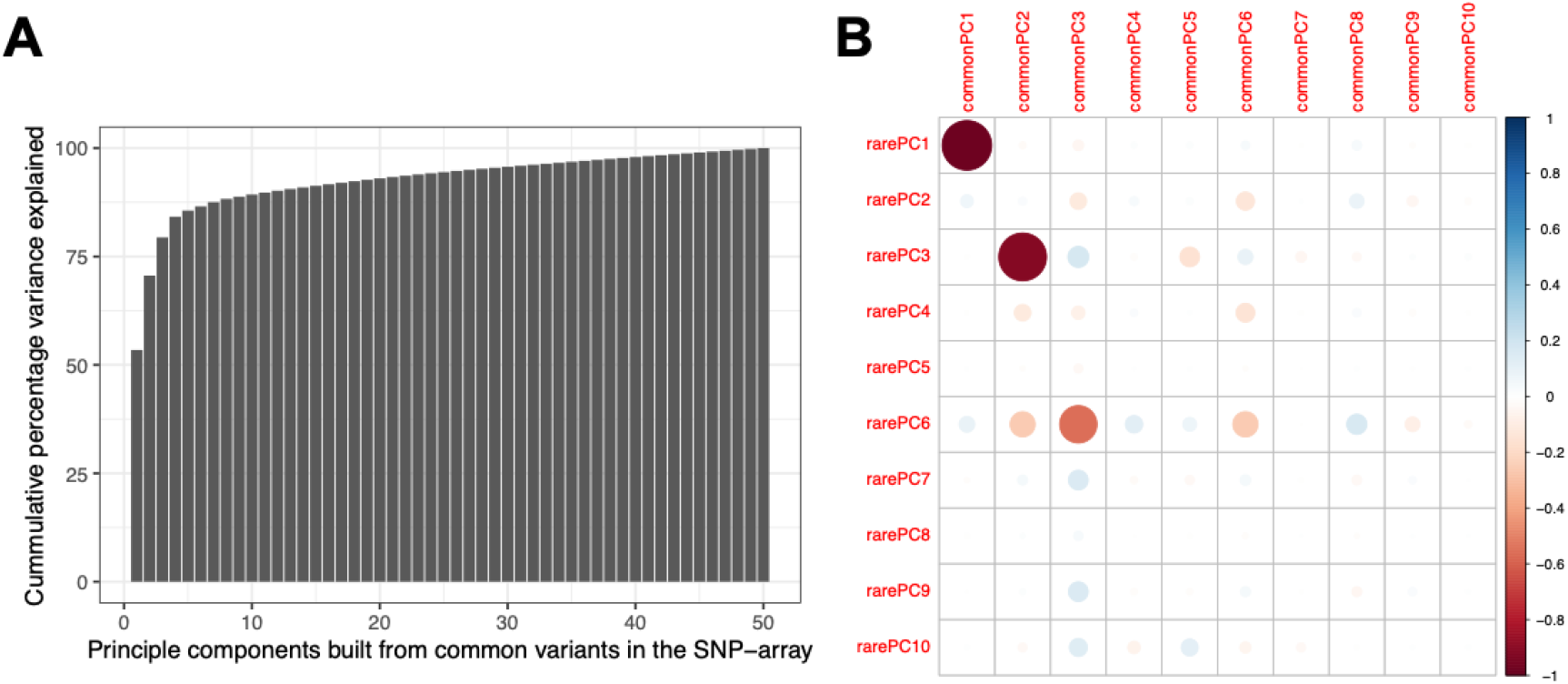
Principal component analyses of SNP-array data. A: The cumulative percentage variance explained across the 50PCs generated from common variants in the SNP-array. B: The correlation plot of the first 10 PCs generated by common variants compared to the first 10 PCs generated by rare variants.

Previous work in the lab by ^75^ demonstrated that increased runs of homozygosity (ROHs) in the genome was associated with several conditions such as anxiety and type 2 diabetes. We sought to explore if the fraction of the genome in ROHs (F_ROH_) is a possible confounder for these recessive findings, by adding it as a covariate and rerunning the association tests for the 42 lead SNPs in the WES5Kimputation dataset. ROHs were calledby PLINK1.9 on the SNP-array data with the following specifications: maximum inverse density 50kb/SNP, maximum internal gap 1000kb, minimum SNP count 50, maximum 1 heterozygous in scanning window hit, maximum 4 missing calls in scanning window hit and a scanning window sizeof 50. The total length of ROHs (in kb) was then divided by the length of the autosome (approximately 2700000kb) to obtain F_ROH_. After controlling for F_ROH_, the recessive p-values correlate well with those obtained without controlling for it, suggesting that this additional covariate is unnecessary (Figure S11).

**Figure S11.**
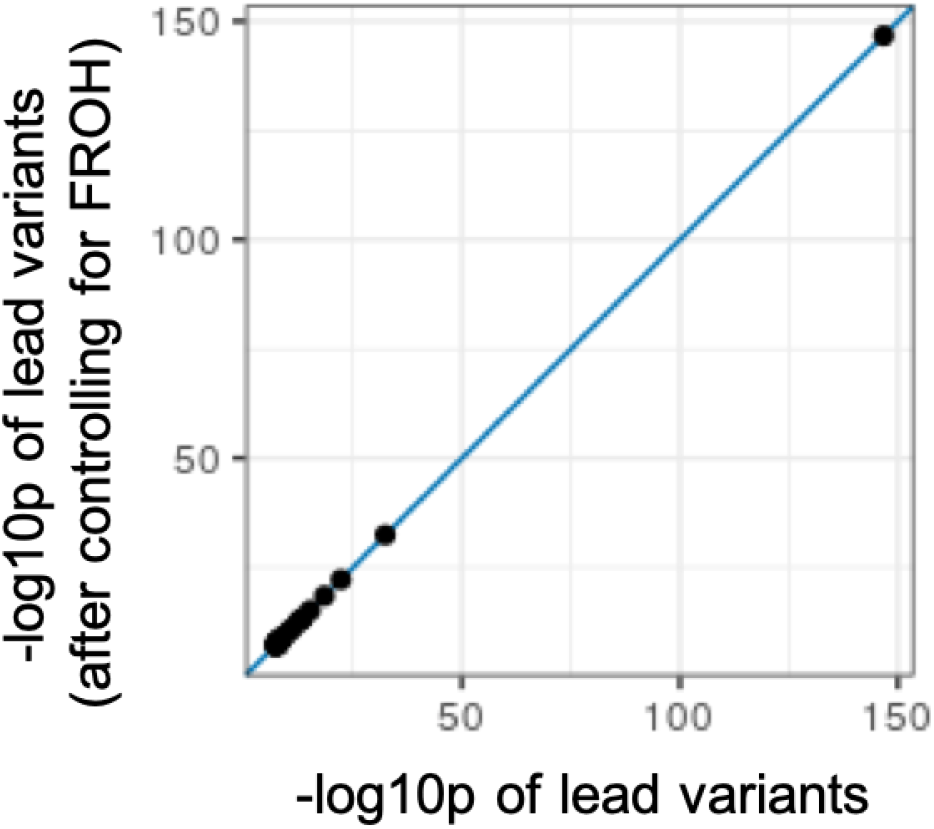
Scatter plot of the WES5Kimputation lead variants p-values (-log10p) compared to their p-values when the recessive test is rerun to control for ^F^ROH.

## Appendix-8: Characteristics of loci identified as significant in the recessive association testing

Information about the 185 lead variants passing p<5×10^−8^ are tabulated in Table S5. The frequency and consequence distributions of the lead variants are shown in Figure S12. Despite applying an exome reference panel including many rare protein-coding variants, many of the findings from the WES5K imputation were still common and intronic, implicating variants near exonic regions that happened to be captured with WES, or common SNPs from the GSA backbone. Similarly, the majority of the significant hits from the whole genome TOPMEDimputation were within the non-coding regions. This is expected as common variants have better power and 99% of the human genome is non-coding.

**Figure S12.**
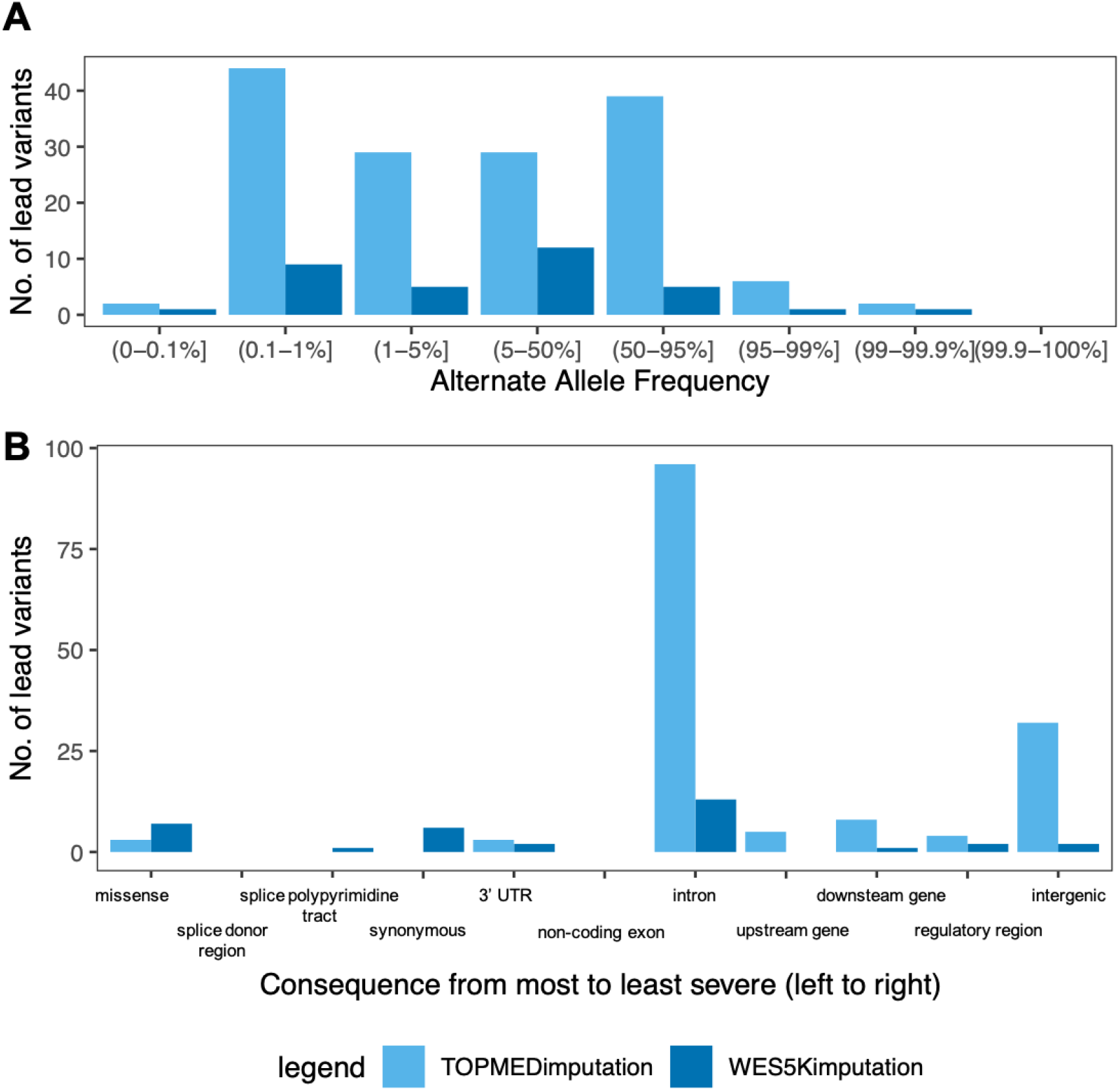
Characteristics of lead SNPs in the recessive findings. A: Distribution of the allele frequencies of the lead SNPs. B: Distribution of the variant consequences of the lead SNPs.

As an example for plotting and visualisation purposes, we plotted the recessive tests performed between the TOPMEDImputation and D58[Other hereditary haemolytic anaemias]. The Quantile-quantile (QQ) plot (Figure-S13A) suggests that the tests are underpowered, as we see substantial deflation of the test statistics below what is expected under the null (lambda = 0.56). When we split the variants contributing to these tests into common (AF > 5%) and low frequency variants (AF ≤ 5%), indeed we see that the lambda of 0.94 for the common variants is close to 1 (Figure-S13BC), while the lambda for the rare variants is low at 0.21 (Figure-S13C), indicating that the deflation is likely due to reduced power for rare variants.

**Figure S13.**
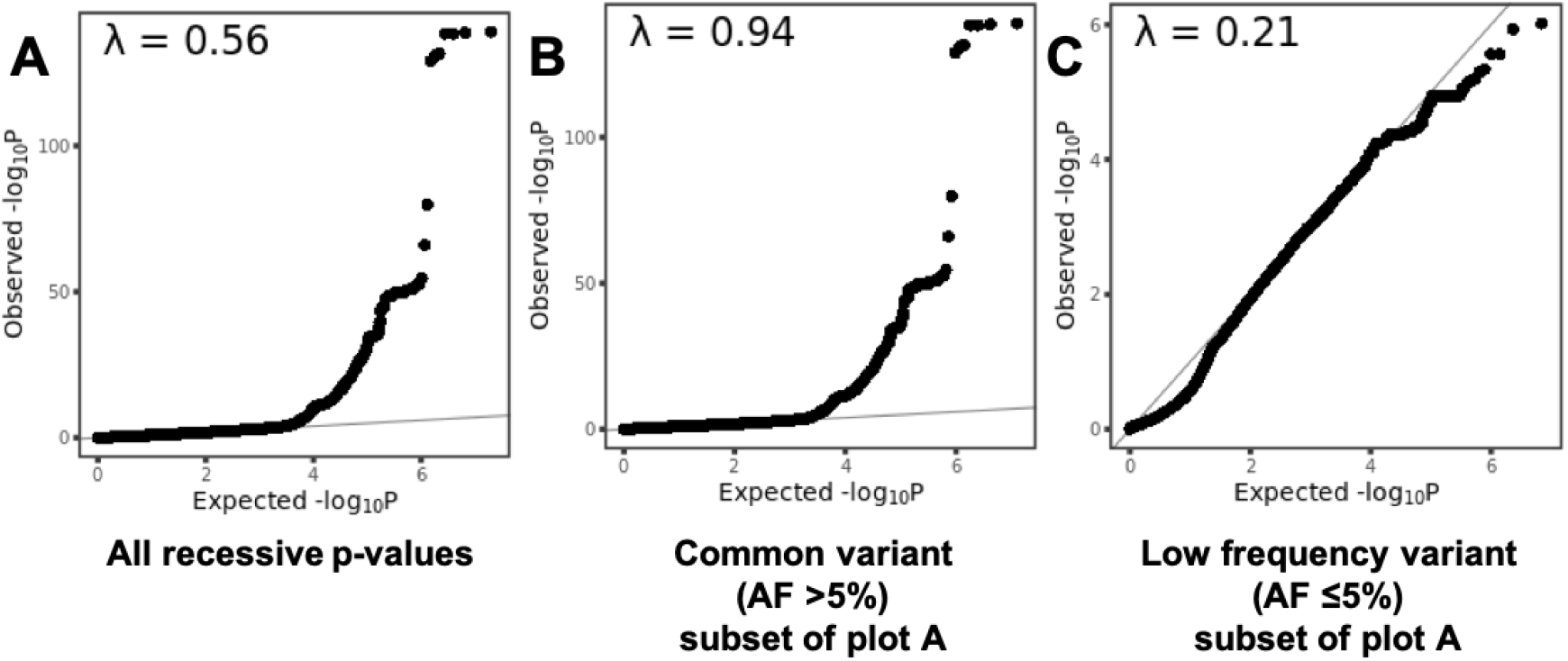
Quantile-quantile (QQ) plots (B-D) for the phenotypes that have Bonferroni-significant findings. In A, all the recessive p-values in the D58[Other hereditary haemolytic anaemias] run have been plotted. We then split the QQ-plot into common variants (AF > 5%, B) and low frequency variants (C).

## Appendix-9: Evaluating the dominance deviation of the 185 recessive findings

For the lead variants of the 185 recessive loci, 152 lead variants were not GWS in the additive test. Looking at the distribution of AFs, recessive hits that were GWS in the additive test had higher AFs than hits that were insignificant in the additive test, suggesting that common variants simply had more power to be detected under the additive model even if the underlying pattern of inheritance might be recessive. (Figure-S14A).

For the lead variants of the significant recessive associations, we also reran the additive test (step 2 of REGENIE) after removing homozygous individuals, to explore heterozygous effects. When doing this, all but two lead variants had p-values below GWS, demonstrating both the weight of these homozygotes on the significant results in the original additive tests and the loss of power by reducing the sample size of the tests. The remaining two lead variants likely have strong heterozygous effects that can be detected even at reduced power. (Figure-S14D)

It is possible that the tests that dropped below GWS after homozygotes were removed may still have heterozygous effects. We compared the betas in the different models of testing. As expected, additive tests estimated betas that were smaller in magnitude compared to the recessive tests (Figure-2B). After dropping the homozygotes, 107 of these tests became insignificant (p-value > 0.05), but for tests that remained nominally significant, their betas correlated well with the full additive tests, though again, their estimated effect sizes were smaller (Figure-S14E). These nominally-significant additive tests performed without homozygous individuals demonstrate heterozygous effects despite reduced power.

When we removed the homozygous individuals for the additive tests, a small minority of tests (18/185 lead variants) could not be run in REGENIE when the homozygotes were excluded. These tests tend to have higher AFs greater than 0.7 (suggesting that the removal of homozygotes resulted in the test being too underpowered) (Figure-S14B).

**Figure S14.**
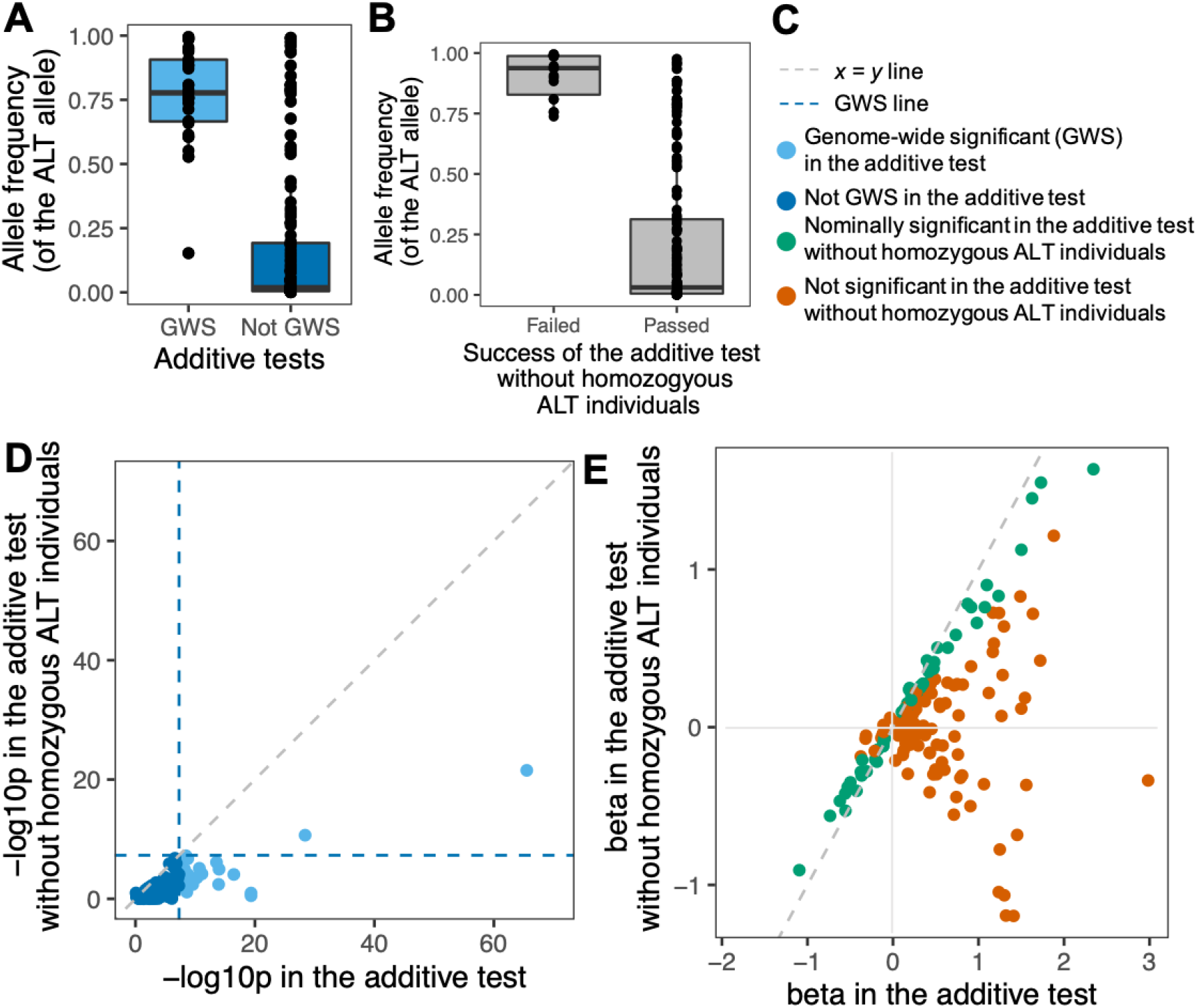
Exploring the recessive lead variants with different models of testing in REGENIE. A: AF distribution of recessive hits that were GWS and not GWS in additive tests. B: AF distribution of recessive hits that could and could not run under the additive model without homozygous individuals. C: legend for the figure. D, E: P-values (-log10p, D) and betas (E) in the additive tests compared to the additive tests without homozygotes.

**Table S6.**
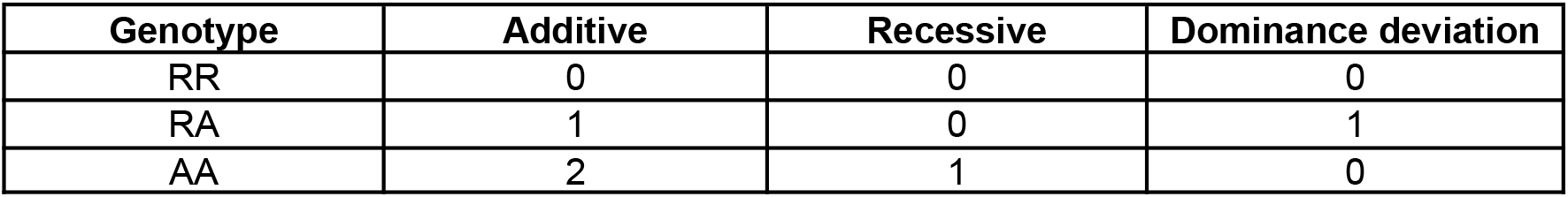
Genotype encodings for the additive, recessive and dominance deviation tests performed as logistic regression tests in R. “RR” refers to the homozygous wild type, “RA” the heterozygous genotype, and “AA” the homozygous alternate genotype.

To further explore whether the recessive model was indeed the best fit for the variants that were significant on the recessive test in REGENIE, we performed logistic regression testing in R, using a genotypic model that included a dominance deviation encoding. For comparison, we also fitted a standard additive and recessive model in R. We re-coded the genotypes to perform additive, recessive and genotypic tests with 2 degrees of freedom, as shown in Table S6 and the equations below.

Additive test:

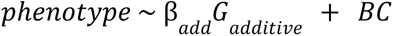

where *G_additive_* is the genotype of the SNP using the additive encoding (0/1/2), β*_add_* is the effect size under an additive model, C is a matrix of covariates (defined below) and B is a vector of effect sizes for those covariates.

Recessive test:

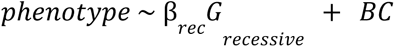

where *G_recessive_* is the genotype of the SNP using the recessive encoding (0/0/1), and β*_rec_* is the effect size under a recessive model.

Genotypic (2 degrees of freedom) test, to extraction dominance deviation:

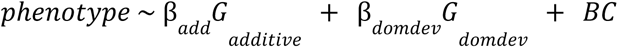

where G*_domdev_* is the genotype of the SNP using the dominance deviation encoding (0/1/0), and β*_domdev_* is the effect size of the dominance deviation under the genotypic model.

The tests were performed on the full cohort (i.e. the set of individuals used in REGENIE, sample sizes in Table S4), as well as on the subset of individuals genetically-inferred to be unrelated by KING (26,579 individuals in the largest cohort, subsetted accordingly depending on the imputation and phenotype tested).

### Covariates

The covariates included were age, sex, age^2^, age x sex, age^2^ x sex and the first ten PCs.

In Figure S15, the results from the additive and recessive tests in R were plotted similarly to those in REGENIE in Figure-2 and gave similar conclusions to those noted earlier in the main text.

**Figure S15.**
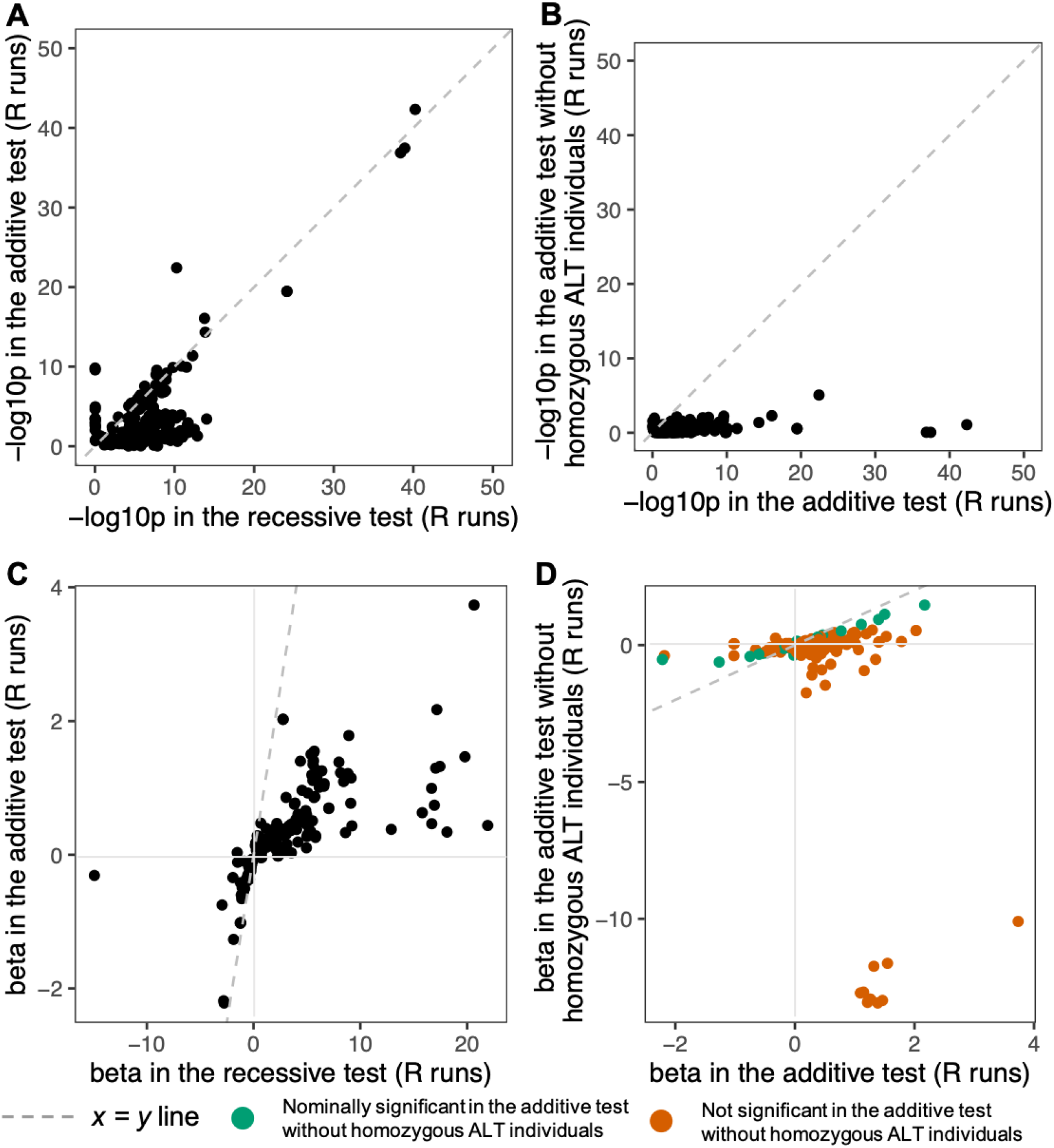
Exploring the recessive lead variants with different models of testing in R. A,C: P-values (-log10p, A) and betas (C) in the recessive tests compared to the additive tests in R. B,D: P-values (-log10p, B) and betas (D) in the additive tests compared to the additive tests without homozygotes in R.

Before considering the results from the dominance deviation tests, we compared the results from the standard recessive model between R logistic regression and REGENIE. We found that the -log10(p-values) correlated well (Figure-S16A, linear regression slope = 1.1, R^2^ = 0.96). This correlation still held when restricting to a set of 26,579 unrelated individuals (in the largest set) in R, though unsurprisingly there was less power (linear regression slope = 0.6, R^2^ = 0.93). Still, there were some REGENIE tests that were not significant (p-value > 0.05) in the R logistic regression, and these outliers had much larger effect size estimates in R (Figure-S16B). Otherwise, for tests that were nominally significant in R, their betas correlated well with REGENIE (linear regression slope = 0.98, R^2^ = 0.98). The outlier R tests that had insignificant p-values tended to involve rarer variants (Figure-S16C, Wilcoxon two-sided p-value = 2×10^−4^), although the distribution of phenotype case counts were similar (Figure-S16C, Wilcoxon two-sided p-value = 0.27). They were excluded from subsequent analyses described below.

When fitting the genotypic model, 76% (or 140) of these lead variants had nominally significant dominance deviation p-values. These variants tended to be at least nominally significant in the R recessive test as well (chi-square test p-value = 1.3×10^−11^, Figure-S16D). For the tests that had an insignificant dominance deviation p-value, we cannot rule out that the underlying inheritance pattern may still be recessive, especially as some also had recessive tests with insignificant p-values in R, suggesting that R is an imperfect model to replicate the results from the more complex model fitted by REGENIE.

Regardless, as expected, the recessive and dominance deviation p-values were correlated (linear regression of their log10-transformed p-values: slope = 4, R^2^ = 0.3, p-value <2.2×10^−16^) (Figure-S16E), with hits having a nominally significant dominance deviation tending to have lower recessive p-values (Wilcoxon two-sided p-value = 2.8×10^−5^). We also saw that hits having a nominally significant dominance deviation also tended to have a larger difference between their recessive and additive p-values (Wilcoxon two-sided p-value = 3×10^−9^) (Figure-S16F). We did not find any difference in the recessive betas, distribution of AFs and case counts between the hits with nominally significant and insignificant dominance deviation p-values (Wilcoxon two-sided p-value 0.3, 0.7 and 0.2 respectively, Figure-S16G-H).

In summary, from removing homozygotes and re-performing the additive tests, we demonstrated that several of our recessive findings may harbour mild heterozygous effects. In addition, fitting the genotypic model in R provided further support for at least three-quarters of our findings being truly recessive.

**Figure S16.**
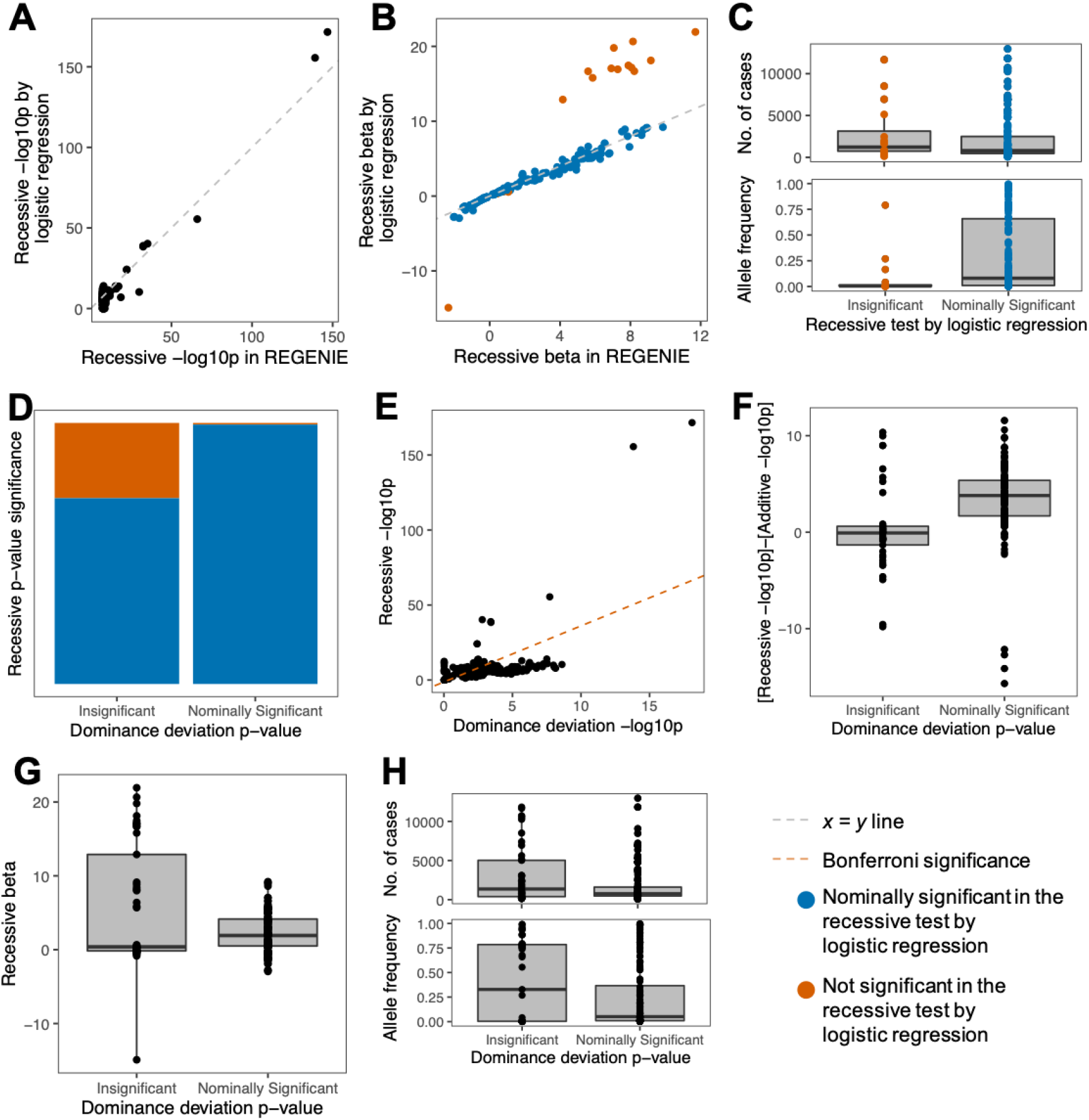
Logistic regression testing in R for the recessive findings. A,B: P-values (-log10p, A) and betas (B) in REGENIE compared to logistic regression testing. C: Distributions of AF and case counts between nominally significant and insignificant (p-value > 0.05) tests in the recessive logistic regression. D-F: Distribution of recessive logistic regression p-values(D-E), the differences between the recessive -log10p and the additive -log10p (F), betas (G), and AF and case counts (H) across the dominance deviation tests.

## Appendix-10: Replication in other cohorts

Table S7 and Table S8 list the Genes & Health phenotypes being matched to FinnGen phenotypes and GERA phenotypes respectively.

The current locus definition (r^2^>0.25 and within 1.5Mb of the lead variant) was chosen so as not to inflate the pairs of independent findings we report, but for the purpose of replication in other cohorts, they are not as stringent as the one described in Huang et al. 2022. For completeness, we repeated the replication calculations with the Huang et al. 2022 cutoffs, so as to calculate a PAT in a manner closer to what was described in the literature. For each significant locus in G&H, we first identified proxy variants as variants that are within a +/-50kb window from the lead variant with LD R^2^ ≥0.8, and a p-value ≤100 times the p-value of the lead variant. The PAT in FinnGen reduced from 23% to 21% for genome-wide significant loci with this more stringent locus definition.

## Appendix-11: Single variant associations with phenotypes found to be significantly associated with genome-wide homozygosity (F_ROH_) in Malawsky et al. (2023)

**Table S9.** Single recessive associations involving phenotypes found to be significantly associated with genome-wide homozygosity in Malawsky et al. (2023) ^18^.

**homozygosity ( $F_{ROH}$ ) in Malawsky et al. (2023)**
| Gene | Lead variant | AF | CSQ | Phenotype | OR | p-value | DD<br>p-value |
| --- | --- | --- | --- | --- | --- | --- | --- |
| **$F_{ROH}$-Associated Phenotypes:** |  |  |  |  |  |  |  |
| **A09[Other gastroenteritis and colitis of infectious and unspecified origin]** |  |  |  |  |  |  |  |
| . | 2:171680912:G:A | 0.3 | intergenic | A09[Other gastroenteritis and colitis of infectious and unspecified origin] | 0.6 | 2.7E-08 | 6.0E-02 |
| **E11[Type 2 diabetes mellitus]** |  |  |  |  |  |  |  |
| . | 6:104459545:G:A | 0.01 | regulatory region | E11[Type 2 diabetes mellitus] | 38 | 3.6E-08 | 7.2E-06 |
| Y_RNA | 10:92706738:G:C | 0.7 | downstream gene | GNH0242 Type 2 Diabetes narrow | 0.8 | 8.8E-09 | 1.0E-01 |
| Y_RNA | 10:92706682:A:G | 0.8 | downstream gene | Type 2 Diabetes | 0.8 | 4.3E-09 | 5.6E-02 |
| ADAMTS16 | 5:5261658:G:A | 0.01 | intron | E14[Unspecified diabetes mellitus] | 296 | 5.1E-09 | 6.9E-06 |
| UBE2E2 | 3:23530638:C:A | 0.001 | intron | E14[Unspecified diabetes mellitus] | 2664 | 4.6E-08 | 9.6E-01 |
| SNX5 | 20:17949555:G:T | 0.01 | intron | GNH0244 Unspecified or Rare Diabetes narrow | 63 | 1.4E-08 | 3.5E-03 |
| **E78[Disorders of lipoprotein metabolism and other lipidaemias]** |  |  |  |  |  |  |  |
| APOA5 | 11:116796367:A:G | 0.9 | upstream gene | E78[Disorders of lipoprotein metabolism and other lipidaemias] | 0.8 | 3.3E-11 | 7.1E-03 |
| **F41[Other anxiety disorders]** |  |  |  |  |  |  |  |
| . | 2:236834949:C:T | 0.01 | regulatory region | F40[Phobic anxiety disorders] | 138 | 2.7E-08 | 2.6E-03 |
| . | 7:131782270:G:A | 0.08 | intergenic | Anxiety and phobia | 1.8 | 3.5E-08 | 4.8E-03 |
| . | 7:131780486:G:A | 0.08 | intergenic | F41[Other anxiety disorders] | 1.9 | 7.7E-10 | 2.7E-03 |
| . | 7:131773247:A:G | 0.1 | regulatory region | F41[Other anxiety disorders] | 1.8 | 1.8E-08 | 7.7E-05 |
| **H61[Other disorders of external ear]** |  |  |  |  |  |  |  |
| ESRRG | 1:216911690:AG:A | 0.01 | intron | H60[Otitis externa] | 54 | 2.5E-09 | 4.7E-04 |
| **J11[Influenza, virus not identified]** |  |  |  |  |  |  |  |
| PKHD1 | 6:52012065:G:GT | 0.004 | intron | J12[Viral pneumonia, not elsewhere classified] | 271 | 3.1E-09 | 6.6E-06 |
| PKHD1 | 6:52012065:G:GT | 0.004 | intron | B97[Viral agents as the cause of diseases classified to other chapters] | 180 | 2.7E-09 | 1.9E-05 |
| . | 6:51529735:A:G | 0.002 | intergenic | B97[Viral agents as the cause of diseases classified to other chapters] | 430 | 2.4E-08 | 8.0E-04 |
| **J34[Other disorders of nose and nasal sinuses]** |  |  |  |  |  |  |  |
| *ADAMTS 9-AS2* | 3:64802433:A:G | 0.03 | intron | J34[Other disorders of nose and nasal sinuses] | 8 | 1.5E-08 | 1.8E-07 |
| . | 7:48916122:T:C | 0.02 | intergenic | J34[Other disorders of nose and nasal sinuses] | 13 | 5.0E-09 | 1.5E-03 |
| . | 5:91760313:C:G | 0.01 | intron | J34[Other disorders of nose and nasal sinuses] | 11 | 2.4E-08 | 1.5E-03 |
| **L30[Other dermatitis]** |  |  |  |  |  |  |  |
| *PTPRN2* | 7:157970755:T:G | 0.005 | intron | L20[Atopic dermatitis] | 64 | 1.1E-08 | 8.6E-01 |
| *NKAIN1* | 1:31215264:T:C | 0.9 | intron | L21[Seborrhoeic dermatitis] | 1.3 | 6.2E-09 | 8.4E-01 |
| *BCAR3* | 1:93847011:TCGG GCGCGGCGG:* | 0.4 | intron | Dermatitis (atopic, contact, other, unspecified) | 1.2 | 1.6E-08 | 6.3E-08 |

## Appendix-12: FinnGen information, ethics statement, materials and methods

FinnGen was launched in 2017 (https://www.finngen.fi/), and it is a pre-competitive collaboration between biobanks in Finland and their supporting organisations such as universities and university hospitals. There is also involvement from international partners from the pharmaceutical industry, and the Finnish biobank cooperative (FINBB). All the FinnGen partners are listed here: https://www.finngen.fi/en/partners.

Patients and control subjects in FinnGen provided informed consent for biobank research, based on the Finnish Biobank Act. Alternatively, separate research cohorts, collected prior the Finnish Biobank Act came into effect (in September 2013) and start of FinnGen (August 2017), were collected based on study-specific consents and later transferred to the Finnish biobanks after approval by Fimea (Finnish Medicines Agency), the National Supervisory Authority for Welfare and Health. Recruitment protocols followed the biobank protocols approved by Fimea. The Coordinating Ethics Committee of the Hospital District of Helsinki and Uusimaa (HUS) statement number for the FinnGen study is Nr HUS/990/2017.

The FinnGen study is approved by Finnish Institute for Health and Welfare (permit numbers: THL/2031/6.02.00/2017, THL/1101/5.05.00/2017, THL/341/6.02.00/2018, THL/2222/6.02.00/2018, THL/283/6.02.00/2019, THL/1721/5.05.00/2019 and THL/1524/5.05.00/2020), Digital and population data service agency (permit numbers: VRK43431/2017-3, VRK/6909/2018-3, VRK/4415/2019-3), the Social Insurance Institution (permit numbers: KELA 58/522/2017, KELA 131/522/2018, KELA 70/522/2019, KELA 98/522/2019, KELA 134/522/2019, KELA 138/522/2019, KELA 2/522/2020, KELA 16/522/2020), Findata permit numbers THL/2364/14.02/2020, THL/4055/14.06.00/2020, THL/3433/14.06.00/2020, THL/4432/14.06/2020, THL/5189/14.06/2020, THL/5894/14.06.00/2020, THL/6619/14.06.00/2020, THL/209/14.06.00/2021, THL/688/14.06.00/2021, THL/1284/14.06.00/2021, THL/1965/14.06.00/2021, THL/5546/14.02.00/2020, THL/2658/14.06.00/2021, THL/4235/14.06.00/2021, Statistics Finland (permit numbers: TK-53-1041-17 and TK/143/07.03.00/2020 (earlier TK-53-90-20) TK/1735/07.03.00/2021, TK/3112/07.03.00/2021) and Finnish Registry for Kidney Diseases permission/extract from the meeting minutes on 4th July 2019.

The Biobank Access Decisions for FinnGen samples and data utilized in FinnGen Data Freeze 10 include: THL Biobank BB2017_55, BB2017_111, BB2018_19, BB_2018_34, BB_2018_67, BB2018_71, BB2019_7, BB2019_8, BB2019_26, BB2020_1, BB2021_65, Finnish Red Cross Blood Service Biobank 7.12.2017, Helsinki Biobank HUS/359/2017, HUS/248/2020, HUS/150/2022 § 12, §13, §14, §15, §16, §17, §18, and §23, Auria Biobank AB17-5154 and amendment #1 (August 17 2020) and amendments BB_2021-0140, BB_2021-0156 (August 26 2021, Feb 2 2022), BB_2021-0169,

BB_2021-0179, BB_2021-0161, AB20-5926 and amendment #1 (April 23 2020)and it’s modification (Sep 22 2021), Biobank Borealis of Northern Finland_2017_1013, 2021_5010, 2021_5018, 2021_5015, 2021_5023, 2021_5017, 2022_6001, Biobank of Eastern Finland 1186/2018 and amendment 22 § /2020, 53§/2021, 13§/2022, 14§/2022, 15§/2022, Finnish Clinical Biobank Tampere MH0004 and amendments (21.02.2020 & 06.10.2020), §8/2021, §9/2022, §10/2022, §12/2022, §20/2022, §21/2022, §22/2022, §23/2022, Central Finland Biobank 1-2017, and Terveystalo Biobank STB 2018001 and amendment 25th Aug 2020, Finnish Hematological Registry and Clinical Biobank decision 18th June 2021, Arctic biobank P0844: ARC_2021_1001.

## Supplementary Documents

Tables S5, S7 and S8 can be found here:

https://docs.google.com/spreadsheets/d/1480luaKK33BuwIk3fkq-1Pqyr10bk7Cg/edit?usp=sharing&ouid=101119527407856944679&rtpof=true&sd=true

Table S10 can be found here:

https://docs.google.com/spreadsheets/d/1DArZ5tn_KZABL3ZJHkstHCe_87GPSzz6/edit?usp=sharing&ouid=101119527407856944679&rtpof=true&sd=true

## Acknowledgements

We thank the Human Genetics Informatics team at the Wellcome Sanger Institute for support with variant annotations.

This research was funded in part by Wellcome (grant no. 220540/Z/20/A, “Wellcome Sanger Institute Quinquennial Review 2021–2026”). For the purpose of open access, the authors have applied a CC-BY public copyright licence to any author accepted manuscript version arising from this submission.

Genes & Health is/has recently been core-funded by Wellcome (WT102627, WT210561), the Medical Research Council (UK) (M009017, MR/X009777/1, MR/X009920/1), Higher Education Funding Council for England Catalyst, Barts Charity (845/1796), Health Data Research UK (for London substantive site), and research delivery support from the NHS National Institute for Health Research Clinical Research Network (North Thames). Genes & Health is/has recently been funded by Alnylam Pharmaceuticals, Genomics PLC; and a Life Sciences Industry Consortium of Astra Zeneca PLC, Bristol-Myers Squibb Company, GlaxoSmithKline Research and Development Limited, Maze Therapeutics Inc, Merck Sharp & Dohme LLC, Novo Nordisk A/S, Pfizer Inc, Takeda Development Centre Americas Inc.

T. H. Heng is supported by the Agency for Science, Technology, and Research (A∗STAR) National Science Scholarship.

We thank Social Action for Health, Centre of The Cell, members of our Community Advisory Group, and staff who have recruited and collected data from volunteers. We thank the NIHR National Biosample Centre (UK Biocentre), the Social Genetic & Developmental Psychiatry Centre (King’s College London), Wellcome Sanger Institute, and Broad Institute for sample processing, genotyping, sequencing and variant annotation.

This work uses data provided by patients and collected by the NHS as part of their care and support.

We thank: Barts Health NHS Trust, NHS Clinical Commissioning Groups (City and Hackney, Waltham Forest, Tower Hamlets, Newham, Redbridge, Havering, Barking and Dagenham), East London NHS Foundation Trust, Bradford Teaching Hospitals NHS Foundation Trust, Public Health England (especially David Wyllie), Discovery Data Service/Endeavour Health Charitable Trust (especially David Stables), Voror Health Technologies Ltd (especially Sophie Don), NHS England (for what was NHS Digital) - for GDPR-compliant data sharing backed by individual written informed consent.

We want to acknowledge the participants and investigators of the FinnGen study. The FinnGen project is funded by two grants from Business Finland (HUS 4685/31/2016 and UH 4386/31/2016) and the following industry partners: AbbVie Inc., AstraZeneca UK Ltd, Biogen MA Inc., Bristol Myers Squibb (and Celgene Corporation & Celgene International II Sàrl), Genentech Inc., Merck Sharp & Dohme LCC, Pfizer Inc., GlaxoSmithKline Intellectual Property Development Ltd., Sanofi US Services Inc., Maze Therapeutics Inc., Janssen Biotech Inc, Novartis Pharma AG, and Boehringer Ingelheim International GmbH. Following biobanks are acknowledged for delivering biobank samples to FinnGen: Auria Biobank (www.auria.fi/biopankki), THL Biobank (www.thl.fi/biobank), Helsinki Biobank (www.helsinginbiopankki.fi), Biobank Borealis of Northern Finland (https://www.ppshp.fi/Tutkimus-ja-opetus/Biopankki/Pages/Biobank-Borealis-bri efly-in-English.aspx), Finnish Clinical Biobank Tampere (www.tays.fi/en-US/Research_and_development/Finnish_Clinical_Biobank_Ta mpere), Biobank of Eastern Finland (www.ita-suomenbiopankki.fi/en), Central Finland Biobank (www.ksshp.fi/fi-FI/Potilaalle/Biopankki), Finnish Red Cross Blood Service Biobank (www.veripalvelu.fi/verenluovutus/biopankkitoiminta), Terveystalo Biobank (www.terveystalo.com/fi/Yritystietoa/Terveystalo-Biopankki/Biopankki/) and Arctic Biobank (https://www.oulu.fi/en/university/faculties-and-units/faculty-medicine/northern-fi nland-birth-cohorts-and-arctic-biobank). All Finnish Biobanks are members of BBMRI.fi infrastructure (www.bbmri.fi). Finnish Biobank Cooperative -FINBB (https://finbb.fi/) is the coordinator of BBMRI-ERIC operations in Finland. The Finnish biobank data can be accessed through the Fingenious® services (https://site.fingenious.fi/en/) managed by FINBB. The team of investigators in FinnGen are listed in Supplementary Table S10.

We want to thank the Genes & Health Research Team (in alphabetical order by surname): Shaheen Akhtar, Mohammad Anwar, Omar Asgar, Samina Ashraf, Saeed Bidi, Gerome Breen, James Broster, Raymond Chung, David Collier, Charles J Curtis, Shabana Chaudhary, Grainne Colligan, Panos Deloukas, Ceri Durham, Faiza Durrani, Fabiola Eto, Sarah Finer, Joseph Gafton, Ana Angel, Chris Griffiths, Joanne Harvey, Teng Heng, Sam Hodgson, Qin Qin Huang, Matt Hurles, Karen A Hunt, Shapna Hussain, Kamrul Islam, Vivek Iyer, Benjamin M Jacobs, Georgios Kalantzis, Ahsan Khan, Claudia Langenberg, Cath Lavery, Sang Hyuck Lee, Daniel MacArthur, Sidra Malik, Daniel Malawsky, Hilary Martin, Dan Mason, Rohini Mathur, Mohammed Bodrul Mazid, John McDermott, Caroline Morton, Bill Newman, Elizabeth Owor, Asma Qureshi, Shwetha Ramachandrappa, Mehru Raza, Jessry Russell, Nishat Safa, Miriam Samuel, Moneeza Siddiqui, Michael Simpson, John Solly, Marie Spreckley. Daniel Stow, Michael Taylor, Richard C Trembath, Karen Tricker, David A van Heel, Klaudia Walter, Caroline Winckley, Suzanne Wood, John Wright, Ishevanhu Zengeya, Julia Zöllner.

Most of all we thank all of the volunteers participating in Genes & Health.

## Author contributions

T.H.H. helped plan the project, performed the data analysis and drafted the manuscript. K.W. supervised the association testing. Q.Q.H. supervised the QC and, with D.A.v.H, performed the TOPMED-r2 imputation. J.K. and M.J.D. ran the recessive testing in FinnGen. H.H. advised on FinnGen data and recessive association testing. D.M. contributed to the power and F_ROH_ calculations. G.K. advised on imputation and association testing. D.A. v.H. supervised the collection and curation of G&H data and advised on association testing. H.C.M planned and led the project, and helped draft the manuscript. All authors reviewed the manuscript.

## Data and code availability

G&H data is available for analysis in a secure Trusted Research Environment. Application can be made to the G&H executive: https://www.genesandhealth.org/research/scientists-using-genes-health-scientific-research

Information on how to access FinnGen data can be found here: https://www.finngen.fi/en/access_results

Software used in the data analysis are publicly available and have been cited. Code written to run these algorithms is available upon reasonable request to the authors.

